# Comparative assessment of methods for short-term forecasts of COVID-19 hospital admissions in England at the local level

**DOI:** 10.1101/2021.10.18.21265046

**Authors:** Sophie Meakin, Sam Abbott, Nikos Bosse, James Munday, Hugo Gruson, Joel Hellewell, Katherine Sherratt, CMMID COVID-19 Working Group, Sebastian Funk

## Abstract

**Background:** Forecasting healthcare demand is essential in epidemic settings, both to inform situational awareness and facilitate resource planning. Ideally, forecasts should be robust across time and locations. During the COVID-19 pandemic in England, it is an ongoing concern that demand for hospital care for COVID-19 patients in England will exceed available resources.

**Methods:** We made weekly forecasts of daily COVID-19 hospital admissions for National Health Service (NHS) Trusts in England between August 2020 and April 2021 using three disease-agnostic forecasting models: a mean ensemble of autoregressive time series models, a linear regression model with 7-day-lagged local cases as a predictor, and a scaled convolution of local cases and a delay distribution. We compared their point and probabilistic accuracy to a mean-ensemble of them all, and to a simple baseline model of no change from the last day of admissions. We measured predictive performance using the Weighted Interval Score (WIS) and considered how this changed in different scenarios (the length of the predictive horizon, the date on which the forecast was made, and by location), as well as how much admissions forecasts improved when future cases were known.

**Results:** All models outperformed the baseline in the majority of scenarios. Forecasting accuracy varied by forecast date and location, depending on the trajectory of the outbreak, and all individual models had instances where they were the top- or bottom-ranked model. Forecasts produced by the mean-ensemble were both the most accurate and most consistently accurate forecasts amongst all the models considered. Forecasting accuracy was improved when using future observed, rather than forecast, cases, especially at longer forecast horizons.

**Conclusions:** Assuming no change in current admissions is rarely better than including at least a trend. Using confirmed COVID-19 cases as a predictor can improve admissions forecasts in some scenarios, but this is variable and depends on the ability to make consistently good case forecasts. However, ensemble forecasts can make forecasts that make consistently more accurate forecasts across time and locations. Given minimal requirements on data and computation, our admissions forecasting ensemble could be used to anticipate healthcare needs in future epidemic or pandemic settings.

## Background

The sheer volume of SARS-CoV-2 reported cases in England combined with a substantial case-hospitalisation rate amongst high-risk groups [1,2], has resulted in an extremely high demand for hospital care in England. As such, it is an ongoing concern that demand for hospital care will exceed available resources. This worst-case scenario has seen patients with COVID-19 receiving lower-quality care [3], as well as cancellations of planned surgeries or routine services; in the United Kingdom, the National Health Service (NHS) faced a substantial backlog of patient care throughout the COVID-19 pandemic [4].

Forecasting healthcare requirements during an epidemic is critical for planning and resource allocation [5–7], and short-term forecasts of COVID-19 hospital activity have been widely used during the COVID-19 pandemic to support public health policy (e.g. [8–11]). While national or regional forecasts provide a big-picture summary of the expected trajectory of COVID-19 activity, they can mask spatial heterogeneity that arises through localised interventions or demographic heterogeneity in the risk of exposure or severity [12]. Small-scale forecasts have been used to support local COVID-19 responses (e.g. in Austin, Texas, USA [9]), as well as to forecast non-COVID-19 or more general healthcare demands at the hospital level [13,14]. Forecasts of hospital admissions are also an essential step to forecasting bed or intensive care unit (ICU) demand (e.g. [11,14,15]).

In theory, future admissions are a function of recent cases in the community, the proportion of cases that require and seek health care (the case-hospitalisation rate (CHR)), and the delay from symptom onset to hospital admission. However, forecasting admissions from community cases is challenging as both the CHR and admission delay can vary over time. The CHR depends on testing effort and strategy (how many symptomatic and asymptomatic cases are identified), the age-distribution of cases [1], and the prevalence of other COVID-19 risk factors amongst cases [12]. Retrospective studies of COVID-19 patients reported a mean delay from symptom onset to hospital admission to be 4.6 days in the UK [16] and 5.7 days in Belgium [17], but this varies by age and place of residence (e.g., care-home residents have a longer average admissions delay than non-residents) [17]. Forecasting studies have found that cases are predictive of admissions with a lag of only 4 - 7 days [10,15]. Given the short estimated delay between cases and future admissions, to make short-term forecasts of admissions therefore also requires forecasts of cases. While some studies consider mobility and meteorological predictors with longer lags [15], they lack a direct mechanistic relationship with admissions and may have only a limited benefit. Besides structural challenges, models are subject to constraints of data availability in real-time and at the relevant spatial scale (by hospital or Trust (a small group of hospitals) for admissions, and local authority level for cases and other predictors).

Models need to be sufficiently flexible to capture a potentially wide range of epidemic behaviour across locations and time, but at the same time should produce results sufficiently rapidly to be updated in a reasonable amount of time. Autoregressive time series models are widely used in other forecasting tasks (e.g. [18,19]), including in healthcare settings [13], and scale easily to a large number of locations; however, since forecasts are, in the simplest case, based solely on past admissions, they may not perform well when cases (and admissions) are changing quickly. Predictors can be incorporated into generalised linear models (GLMs) with uncorrelated [13] or correlated errors [15]; for lagged predictors, the lag (or lags) usually needs to be predetermined. Alternatively, admissions can be modelled as a scaled convolution of cases and a delay distribution; this method can also be used to forecast deaths from cases or admissions (e.g. [20]). The forecasting performance of both GLMs and convolution models beyond the shortest forecast horizon will be affected by the quality of the case forecasts (or any other predictors), which may vary over time or across locations.

One way to attempt improving the robustness of forecasts is to combine them into an ensemble forecast, whereby predictions from several different models are combined into a single forecast. This reduces reliance on a single forecasting model and, given a minimum quality of the constituent models, the average performance of ensembles is generally comparable, if not better than, its best constituent models [8,21]. Ensemble methods have been widely used in real-time during the COVID-19 pandemic to leverage the contributions of multiple modelling groups to a single forecasting task [8,22,23], as well as previously during outbreaks of influenza [19,24], Ebola virus disease [25], dengue [26] and Zika [27].

In this paper, we make and evaluate weekly forecasts of daily hospital admissions at the level of NHS Trusts during the period August 2020 - April 2021, including two national lockdowns and the introduction and spread of the Alpha SARS-CoV-2 variant. We assess the forecasting performance of three individual forecasting models and an ensemble of these models, and compare their performance to a naive baseline model that assumes no future change from current admissions. Forecasts are made using publicly available data on hospital admissions (by Trust) and COVID-19 cases (by upper-tier local authority (UTLA), a geographic region of England). For forecasting models that use forecast COVID-19 cases as a predictor, we consider the value of making perfect case forecasts.

## Methods

### Data

The majority of hospitalised COVID-19 cases in England are treated at hospitals run by the NHS. NHS Hospital Trusts are organisational units of NHS England, each comprising a small number of hospitals (typically between one and three) and providing care to a small geographical region or for a specialised function [28].

A confirmed COVID-19 hospital patient is any patient admitted who has recently (in the last 14 days) tested positive for COVID-19 following a polymerase chain reaction (PCR) test, including both new admissions with a known test result and inpatient tests. Data on daily Trust-level COVID-19 hospital activity, including COVID-19 hospital admissions, COVID-19 and non-COVID-19 bed occupancy, are published weekly by NHS England and were accessed via the *covid19*.*nhs*.*data* R package [29].

A confirmed COVID-19 case in England is defined as an individual with at least one confirmed positive test from a PCR, rapid lateral flow tests or loop-mediated isothermal amplification (LAMP) test. Positive rapid lateral flow test results can be confirmed with PCR tests taken within 72 hours; if the PCR test results are negative, these are not reported as cases. Aggregated data by UTLA are published daily on the UK Government dashboard and reported totals include both pillar 1 (tests in healthcare settings and for health and care workers) and pillar 2 (community) tests. These data were accessed via the *covidregionaldata* R package [30].

In England, small-scale COVID-19 cases and hospital admissions are reported on different scales: by UTLA and by Trust, respectively. To use UTLA-level cases to make forecasts of Trust-level hospital admissions, we needed to estimate cases at the Trust level too. We used a many-to-many mapping between UTLAs and NHS Trusts that is based on COVID-19 hospital admissions line list data for England. For each Trust-UTLA pair *(t,u)*, the mapping reports the proportion *p*_*t,u*_ of all COVID-19 hospital admissions from UTLA *u* that were admitted to trust *t*. This proportion is based on all COVID-19 hospital admissions in England that were discharged by 30 September 2020, and is constant (i.e. does not change over time). For details of how this mapping was constructed, see Additional file 1: Section 1. This mapping is available in the R package *covid19*.*nhs*.*data* [29].

We estimate the community pressure of COVID-19 cases on Trust *t* as the expected number of COVID-19 cases associated with Trust *t* defined by the Trust-UTLA mapping (rounded to the nearest integer value):

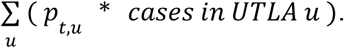

### Trust characteristics

We estimated Trust size as the average total beds available (sum of occupied COVID-19 or non-COVID-19 beds, plus unoccupied beds) from 17 November 2020 to 30 April 2021 (data on non-COVID bed occupancy was not available before 17 November). We calculated total admissions as the sum of all admissions between 01 August 2020 and 30 April 2021 (inclusive). We defined the size of the Trust-UTLA mapping for a Trust as the number of UTLAs matched to each Trust in the probabilistic Trust-UTLA mapping. We measured this with and without a 10% minimum threshold on the proportion of admissions from a UTLA to a Trust, thereby excluding relatively uncommon Trust-UTLA pairs.

To better understand the heterogeneity in Trust-level admissions, we grouped Trusts based on the similarity of their weekly hospital admissions time series. We calculated the pairwise Pearson correlation coefficient between Trusts, excluding Trusts with less than 1000 admissions between August 2020 and April 2021. We then used the complete-linkage clustering algorithm to divide Trusts into seven groups, matching the seven NHS regions in England. In short, the complete-linkage algorithm initially assigns each Trust to its own cluster, then at each step combines the two most similar clusters (as determined by the pairwise correlation), until the desired number of clusters is reached [31]; this is implemented in the *hclust* algorithm in *stats 4*.*1*.*1*.

### Forecasting models

We made weekly forecasts of daily hospital admissions from 04 October 2020 to 25 April 2021 (n = 30 forecast dates). We fitted each of the forecasting models (defined below) independently to each Trust’s unsmoothed and unadjusted Trust-level daily data (past admissions and, where relevant, past estimated cases) on a 6-week rolling window, and made forecasts of future admissions for 1-through 14-day-ahead horizons. We used a rolling, rather than increasing, window on which to fit the models as the local trend in admissions and relationship between cases and admissions was considered likely to change over time. We summarised forecasts as point and predictive quantiles for 1 through 14-day ahead horizons. For models using cases as a predictor of future hospital admissions, we used forecasts of daily UTLA-level COVID-19 cases that were produced and published daily [32].

#### Hospital admissions forecasting models

The motivation for a baseline model is to give a minimum performance threshold that any good model should reasonably exceed. Our baseline model comprised a point (median) forecast equal to the last observed data point for all forecast horizons and Gaussian uncertainty, with standard deviation at horizon *h* equal to 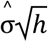, where 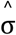 is the standard deviation of the differences of the series [18].

In addition, we used three individual forecasting methods, plus two unweighted ensembles of these three models. The first of the three individual models was a mean-ensemble of three autoregressive time series models (autoregressive integrated moving average (ARIMA), exponential smoothing (ETS) and the baseline defined above) that use only past observed admissions data to forecast future admissions. The second model was a regression model with correlated (ARIMA) errors, with Trust-level cases lagged by *d* days as a predictor. This model uses past observed admissions and past estimated Trust-level cases (estimated via the Trust-UTLA mapping) for forecast horizon *h* ≤ *d*, plus forecast Trust-level cases (again, estimated via the Trust-UTLA mapping) for *h* > *d*, where the optimal value for *d* is chosen by optimising for forecasting performance. The third and final individual model is a convolution of estimated Trust-level cases with the delay from report to admission. This model uses past observed admissions and past and future Trust-level cases. None of the models include a day-of-the-week effect, as it was determined a priori that this was not a consistent feature of the Trust-level data. Further details of the three individual forecasting models can be found in Additional file 1: Section 2 [33–37] (Tables S1 - S4 and Figure S1).

We constructed an unweighted mean-ensemble from the three individual models. The ensemble quantile forecast was made by taking the mean of the quantile forecasts of the individual models at each time point; for example, the mean-ensemble point forecast for a 7-day horizon was the mean of the three individual point forecasts for a 7-day horizon, and the mean-ensemble 90% quantile forecast was the mean of the three individual 90% quantile forecasts.

#### Case forecasting models

The ARIMA regression model and convolution model used COVID-19 cases as a predictor of future hospital admissions, and so we also used forecasts of this quantity. We used daily forecasts of COVID-19 cases by UTLA (n = 174) via estimates and forecasts of the time-varying effective reproduction number, R_t_, accounting for uncertainty in the delay distributions and day-of-the-week effect, produced and published daily [32]; a summary of this approach, henceforth called R_t_ case forecast, is given in Additional file 1: Section 3 [38,39] and full details are given in [40].

The R_t_ case forecasts were occasionally missing due to computational issues or deemed highly improbable due to model errors. As the case forecasts are used as predictors in some of the admissions forecasting models, this could lead to highly improbable (particularly excessively large) admissions forecasts. To address this, we set three criteria by which the R_t_ case forecasts would be replaced by an ARIMA + ETS mean-ensemble time series forecast. We did not expect that this time series ensemble would produce better forecasts than the R_t_ model in all scenarios, but rather that they would be better than missing or implausible forecasts. The three criteria were:

1. The R_t_ case forecast was missing for the UTLA, or
2. The upper bound of the R_t_ case forecast 90% prediction interval exceeded the estimated population size of the UTLA [41], or
3. There was at least one case reported on the forecast date, and the upper bound of the R_t_ case forecast 90% prediction interval exceeded 1000 times the number of cases reported on the forecast date.

We estimated Trust-level case forecasts from the UTLA-level case forecasts using the Trust-UTLA mapping.

### Forecast evaluation

#### Evaluation metrics

We evaluated forecasts against future observed admissions using a number of different metrics that assessed different aspects of point and probabilistic accuracy.

##### Calibration

Calibration assesses the ability of the models to correctly quantify predictive uncertainty. We assessed the calibration of the forecasting models by calculating the empirical coverage: for a forecast horizon, *h*, and prediction interval width, 1 − α, the empirical coverage of a model is calculated as the proportion of forecast targets (across all forecast dates and locations) for which the prediction interval contained the true value; a well-calibrated model has empirical coverage equal to the width of the nominal prediction interval. We calculated the empirical coverage for the 50% and 90% prediction intervals.

##### Sharpness

Sharpness measures the ability of models to make forecasts with narrow (sharp) prediction intervals. We measured sharpness as the weighted sum of the width of the 50% and 90% prediction intervals:

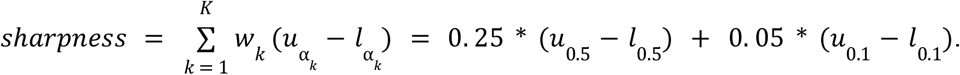

##### Point forecast error

We measured point forecast accuracy with the absolute error (AE) of the median forecast, which is simply the absolute difference between the median forecast, *m*, and the true observed value, *y*: |*m* − *y*|.

##### Probabilistic forecast error

We measured probabilistic forecast accuracy with the weighted interval score (WIS). The WIS is a proper scoring rule, that is, a rule for which a forecaster is incentivised to give their honest forecast to obtain the best score [42]. The WIS comprises a weighted sum of interval scores for quantile forecasts of increasing widths; in this way, the full forecast distribution is summarised in a single value.

The interval score [43] of the central 100(1-α)% predictive interval of forecast *F* is given by

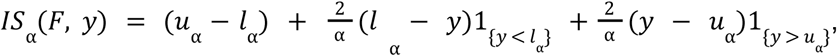

where *l*_α_ and *u*_α_ are the lower and upper bounds of the central 100*(1-α)% interval forecast, *y* is the true observed value, and 1_{·}_ is the indicator function (equal to 1 when the expression inside is true, and 0 otherwise). The first term measures sharpness, and penalises wider interval forecasts; the second term penalises forecasts for overprediction (if the true value, *y*, lies below the lower bound *l*_α_); finally, the third term penalises for underprediction.

Given the point forecast and *K* interval forecasts of width 1 − α_*k*_, *k =* 1,…, *K*, the WIS is then calculated as

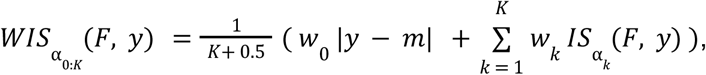

where the standard choice is *w*_*0*_ *=* 1*/2* and 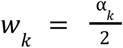 for *k =* 1,…, *K*[43]. In our evaluation, we used *K =* 2 and α _1_ *=* 0. 5, α_2_ = 0. 1, corresponding to the central 50% and 90% prediction intervals, respectively.

To summarise and compare forecast performance in different scenarios (see Section “Forecast comparison” below), we either report the mean value (sharpness and AE), or an adjusted value that does not scale with the number of admissions (WIS). The latter allows us to compare a model’s performance over forecast dates or between Trusts (both of which vary in the number of admissions). Instead of reporting the mean WIS we report two adjusted WIS values: the relative WIS () and the scaled WIS (sWIS), defined as follows using the notation and naming of [21].

First, the pairwise-relative WIS, θ_*A,B*_, for models *A* and *B* is defined as

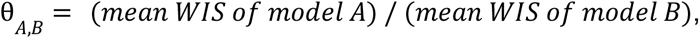

where the mean WIS is the mean in the scenario of interest (e.g. to evaluate models’ overall performance at a 7-day horizon, the mean is taken over all forecast dates and Trusts).

The rWIS for model A, θ_*A*_, is then defined as the geometric mean of the pairwise-relative WIS 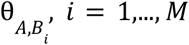, excluding the baseline model. If model A has a smaller relative WIS than model B, then forecasts generated by model A are better than those generated by model B.

The sWIS, 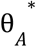, is simply the rWIS normalised by the rWIS for the baseline model:

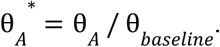

By this definition, the sWIS of the baseline model is always 1, and if 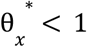 then forecasts produced by model *x* are better than the baseline, and worse if 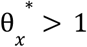.

Forecast evaluation was implemented using the R package *scoringutils* 0.1.7.2 [44].

#### Forecast comparison

We evaluated forecasts made across 7,701 forecast targets, a combination of forecast horizon (7 or 14 days), forecast date (30 total) and Trust (129 until 24 January 2021; then 128 until 14 March 2021; then 127 until the end of April 2021). To fully evaluate model performance, we evaluated forecast performance in the following scenarios:

1. Overall:
  a. By forecast horizon;
  b. By target;
2. By forecast date, split by forecast horizon;
3. By Trust, split by forecast horizon.

In scenario 1a we report empirical coverage by forecast horizon, mean sharpness, mean AE and rWIS. In scenario 1b we simply report the distribution of model rankings over all 7,701 targets as determined by the rWIS. In scenario 2 and 3 we report the mean AE and rWIS by forecast date and Trust, respectively. In all scenarios we choose to report rWIS over the sWIS so that the performance of the baseline model, and how that changes across horizons/dates/locations, can be explicitly included.

#### Value of perfect knowledge of future COVID-19 cases

For models that use forecasted COVID-19 cases to forecast hospital admissions, forecast performance is affected by both the structure of the admissions forecasting model and the quality of the case forecasts (which are made independently of the admissions forecasts and do not form part of this study). Models that use forecast cases to forecast admissions are the ARIMA regression for forecast horizon *h* > *7*, and the convolution model and mean-ensemble for all forecast horizons. To evaluate the performance of the admissions models only, we conducted a retrospective study where the relevant models (noted above) used future observed, rather than forecast, COVID-19 cases to forecast hospital admissions; this represents a best-case scenario for these models, as using future observed cases throughout is equivalent to making a perfect case forecast with no uncertainty.

These retrospective forecasts were scored using the same metrics and scenarios as the real-time forecasts. We also directly compare each model’s performance using observed vs. forecast future COVID-19 cases, where we report the sWIS only.

### Analysis code

Analyses in this paper use the following packages developed by the authors during the COVID-19 pandemic: *covidregionaldata* (version 0.9.2) [30], *covid19*.*nhs*.*data* (0.1.0) [29], *EpiNow2* (1.3.3.8) [20], *EpiSoon* (0.3.0) [36] and *scoringutils* (0.1.7.2) [44]. Fully reproducible code is available at https://github.com/epiforecasts/covid19-hospital-activity.

## Results

### COVID-19 hospital activity August 2020 - January 2021

#### National and regional context

The number of COVID-19 hospital admissions in England was very low at the start of August 2020: during the week 03 - 09 August 2020, national daily admissions ranged between 49 and 78. From early September onwards, admissions began to increase (Figure 1A), predominantly in the Midlands and North of England (Additional file 1: Figure S2). In response to rising cases and admissions, the UK Government introduced a three-tier system of restrictions throughout England on 14 October 2020; by the end of October all major northern cities (including Manchester and Liverpool) were under the strictest Tier 3 measures, and the majority of the rest of the North of England, plus the Midlands, London and parts of Essex were in Tier 2. A national lockdown was introduced from 05 November - 02 December 2020 and during this time admissions fell or plateaued in all NHS regions (Additional file 1: Figure S2). At the end of lockdown (03 December 2020), the majority of local authorities in England re-entered Tier 3, but hospital admissions continued to increase: national daily admissions increased from 1,178 on 02 December 2020, to 1,437 one week later (09 December 2020), and to 1,880 two weeks later (16 December 2020) - already exceeding the early-autumn peak of 1,620 daily admissions. On 19 December 2020 local authorities in the East and South East of England and all London boroughs entered into yet stricter Tier 4 restrictions, and on 06 January 2021 England was placed under the third national lockdown. National daily admissions peaked at 3,895 on 12 January 2021 and subsequently declined throughout January - April 2021. By the end of April, average national daily admissions were fewer than 100 (during the week 19 - 25 April 2021, median = 97, interquartile range (IQR) = 28).

**Figure 1:**
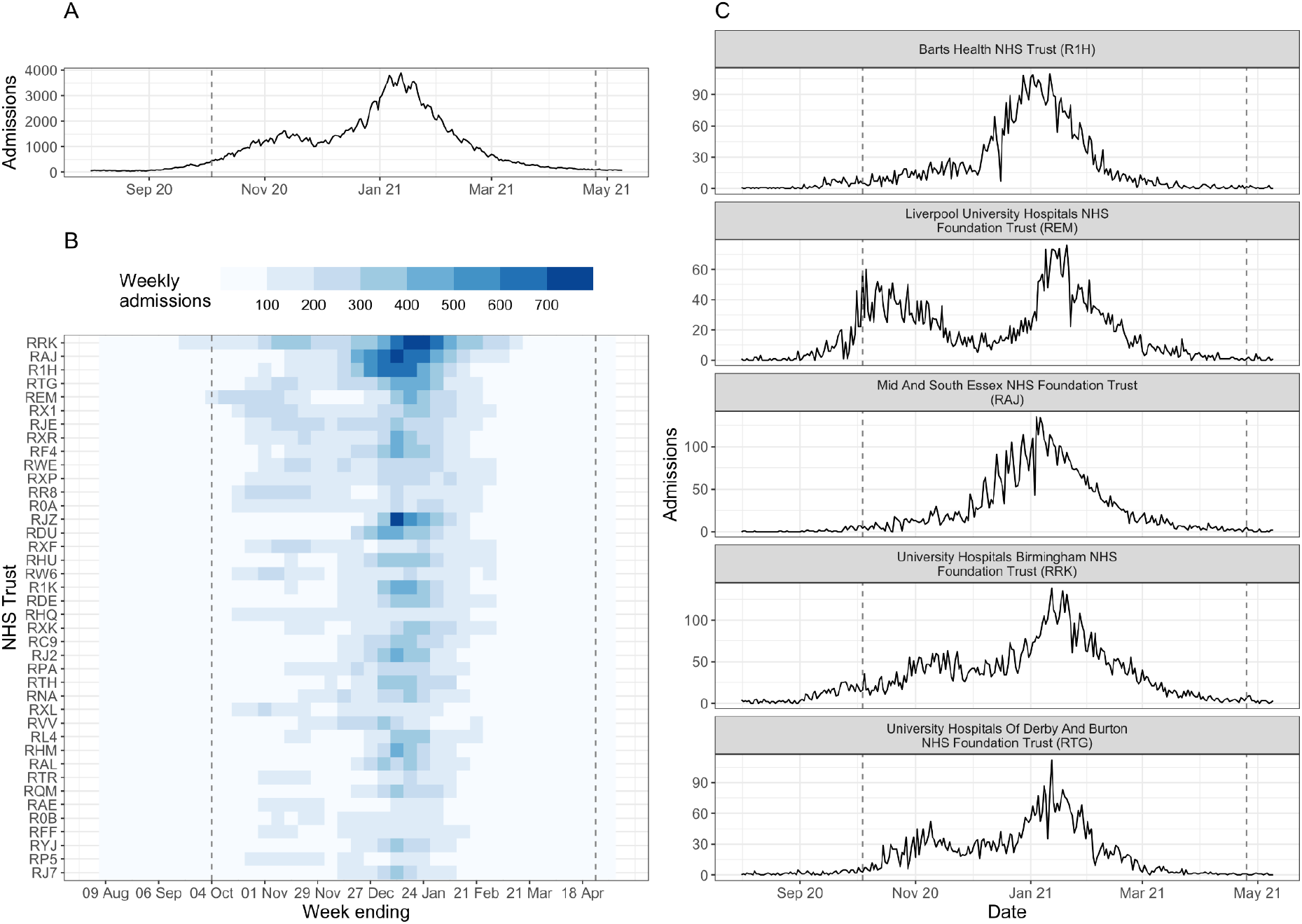
Summary of COVID-19 hospital admissions in England during August 2020 - April 2021. (A) Daily COVID-19 hospital admissions for England. (B) Weekly COVID-19 hospital admissions by NHS Trust (identified by 3-letter code) for the top 40 Trusts by total COVID-19 hospital admissions during August 2020 - April 2021. (C) Daily COVID-19 hospital admissions for top-five Trusts by total COVID-19 hospital admissions. In all panels, the dashed lines denote the date of the first (04 October 2020) and last (25 April 2021) forecast date. Data published by NHS England [45].

Mass vaccination for COVID-19 in England began on 08 December 2020. The rollout was prioritised by age and risk. The initial rollout was amongst care home residents, their carers and individuals aged 80 years and over, then subsequently to all aged 70 years and over and the clinically extremely vulnerable individuals (from 18 January 2021) and then all aged 65 years and over and adults with high-risk underlying health conditions (15 February 2021). By 30 April 2020, 63% and 27% of adults aged 16 and over had received the first and second dose of the vaccine, respectively [46].

#### Trust-level characteristics and hospital admissions

Focusing only on national or regional hospital admissions masks heterogeneity in the trajectory of local-level hospital admissions (Figure 1). Trusts varied in the daily or weekly number of patients admitted, as well as in the occurrence and timing of peaks in admissions (Figure 1B-C). Clusters of Trusts, defined by the pairwise correlation between admissions, clearly show some spatial clustering (reflecting the geographical spread of COVID-19 in England at the time) but are not constrained by the NHS region boundaries (Additional file 1: Figure S3C). Instead, clusters are defined by the occurrence and timing of peaks in admissions (Additional file 1: Figure S3D): for example, cluster 1 includes Trusts in London and the South East of England that had little or no peak in November 2020, while clusters 5-7 comprise Trusts in the North of England where admissions increased earlier and there are two distinct peaks in admissions in November 2020 and January 2021 (Additional file 1: Figure S3C-D). The variation in Trust-level dynamics could be driven by Trust capacity, local COVID-19 case incidence and demography (such as age), pre-existing immunity, and local and/or national restrictions.

Trust-level cases are estimated using the Trust-UTLA mapping. The accuracy of this mapping for a given Trust depends on a number of factors, including: the spatial distribution of cases until 30 September 2020; total admissions to the Trust until 30 September 2020; and the size of the Trust-UTLA mapping. Trusts admit COVID-19 patients from relatively few UTLAs (median = 3, IQR = 2; Additional file 1: Figure S3A), with a small minority of Trusts (typically in London or other large cities such as Birmingham and Manchester) admitting patients from more than 10 UTLAs. Trusts admit the majority of their COVID-19 patients from only 1-2 UTLAs (excluding UTLAs contributing less than 10% of admissions: median = 2, IQR = 1.8).

Estimated total bed capacity and total admissions vary significantly (estimated capacity: median = 579, IQR = 417; total admissions: median = 1,839, IQR = 1,313), and, unsurprisingly, the two are highly correlated (Pearson’s correlation coefficient r = 0.85) (Additional file 1: Figure S3B).

### Forecast evaluation

Additional file 1: Figure S4 shows examples of forecasts made for Manchester University NHS Foundation Trust for the three individual models (time series ensemble, ARIMA regression with 7-day lagged cases as a predictor, and the case-convolution), plus the mean-ensemble of these, and the baseline model of no change.

#### Calibration and sharpness

The empirical coverage of models was generally lower than the nominal coverage of the prediction intervals (Figure 2A and Table S5); the only exception to this is the 50% prediction interval of the time series ensemble, which has empirical coverage of 53% and 54% for a 7- and 14-day horizon, respectively. The ARIMA model has the worst coverage for all forecast horizons as a result of producing overly sharp (narrow) forecasts (sharpness at a 14-day horizon of 0.97, compared to 2.85 for the baseline and 1.57 for the mean-ensemble; Table S5). Although its constituent models are not particularly well-calibrated, the mean-ensemble still has comparatively good empirical coverage: for a 14-day horizon, it has empirical coverage of 0.46 and 0.76 for the 50% and 90% prediction intervals respectively.

**Figure 2:**
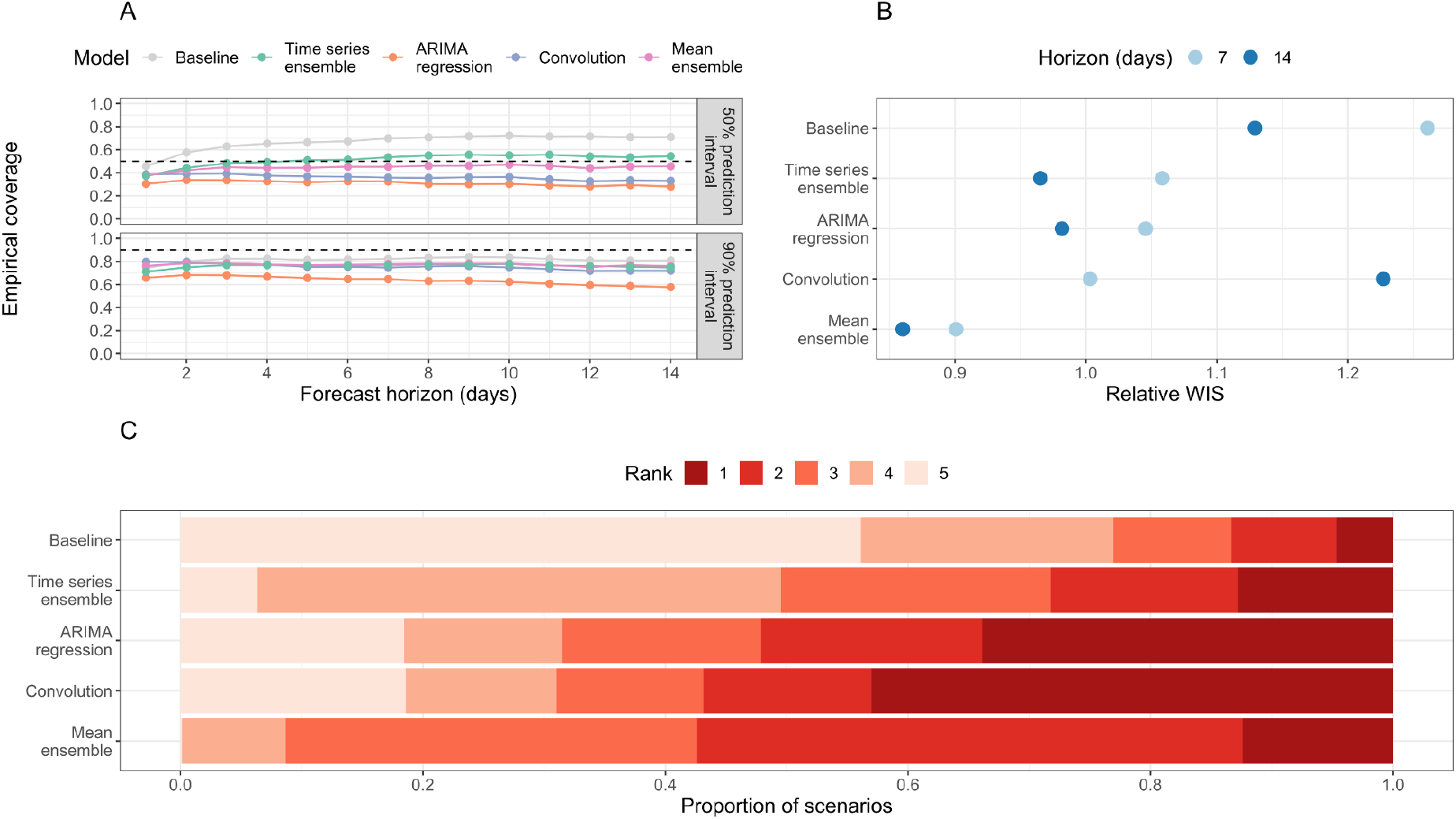
Overall forecasting performance of forecasting models. (A) Empirical coverage of 50% and 90% prediction intervals for 1-14 days forecast horizon. The dashed line indicates the target coverage level (50% or 90%). (B) Relative weighted interval score (rWIS) by forecast horizon (7 and 14 days). (C) Distribution of WIS rankings across all 7,701 targets; for each target, rank 1 is assigned to the model with the lowest relative WIS (rWIS) and rank 5 to the model with the highest rWIS.

#### Overall forecast accuracy

For a 7-day forecast horizon, the time series ensemble and ARIMA regression model use observed data only (hospital admissions, plus confirmed COVID-19 cases in the ARIMA regression model). Both models outperformed the baseline (rWIS = 1.06 and 1.05, respectively, compared to 1.26 for the baseline; Figure 2B, Table S5). The convolution model uses a combination of true and forecast COVID-19 cases, yet was still the best-performing individual model at this horizon (rWIS = 1.00). However, the mean-ensemble clearly outperformed all models and made 29% less probabilistic error than the baseline model (sWIS = 0.90).

For a 14-day forecast horizon, only the time series ensemble uses exclusively observed data (hospital admissions); both the ARIMA regression model and the convolution model use forecast COVID-19 cases. Whilst the relative accuracy of all models decreased (sWIS increases) over a longer horizon (Table S5), the decline in performance was most substantial for the convolution model, which now performed worse than baseline (rWIS = 1.23 compared to 1.13 for the baseline). Despite the worse performance of one of its constituent models, the mean-ensemble still performed well, making 24% less probabilistic error than the baseline model (rWIS = 0.86).

The relative WIS rankings over all 7,701 individual targets showed some variability in forecasting performance (Figure 2C). Interestingly, all individual models (time series ensemble, ARIMA regression and case-convolution) rank first more frequently than the mean-ensemble (in 13%, 34% and 43% of targets, respectively, compared to 12% for the mean-ensemble). However, the mean-ensemble is the most consistent model: it ranks first or second in over half (57%) of targets, and first through third in over 90% of scenarios. In comparison, the individual models often rank fourth or fifth (last). The mean-ensemble also outperforms the baseline in 84% of scenarios, compared to 82% for the time series ensemble, and 75% for both the ARIMA regression and case-convolution models. There are also some targets (approximately 5%) where the baseline outperforms all models (Figure 2C).

#### Forecast accuracy by date

Probabilistic forecasting accuracy and model rankings varied by the date on which forecasts were made (Figure 3 and S5). For a 7-day horizon, the mean-ensemble was the only model to outperform the baseline model (as measured by rWIS) across all forecast dates (Figure 3A). Moreover, the mean-ensemble was the first-ranked (best) model by this metric for 14/30 forecast dates, and was first- or second-ranked for 29/30 dates. The performance of the individual models was more variable. While the time series ensemble outperformed the baseline for 29/30 forecast dates, it was often only the third- or fourth-ranked model (24/30 forecast dates). On the other hand, the convolution model was the top-ranked model for 14/30 forecast dates, but performed particularly poorly on two dates (03 and 10 January 2021; Figure 3A-B). For all models, the biggest improvement in forecasting performance compared to the baseline was at times when hospital admissions were rapidly declining: mid-to-late November 2020 (improvement in rWIS of approximately 30%) and from mid-January 2021 onwards (improvement of up to 49%) (Figure 3A and Additional file 1: Figure S5A).

**Figure 3:**
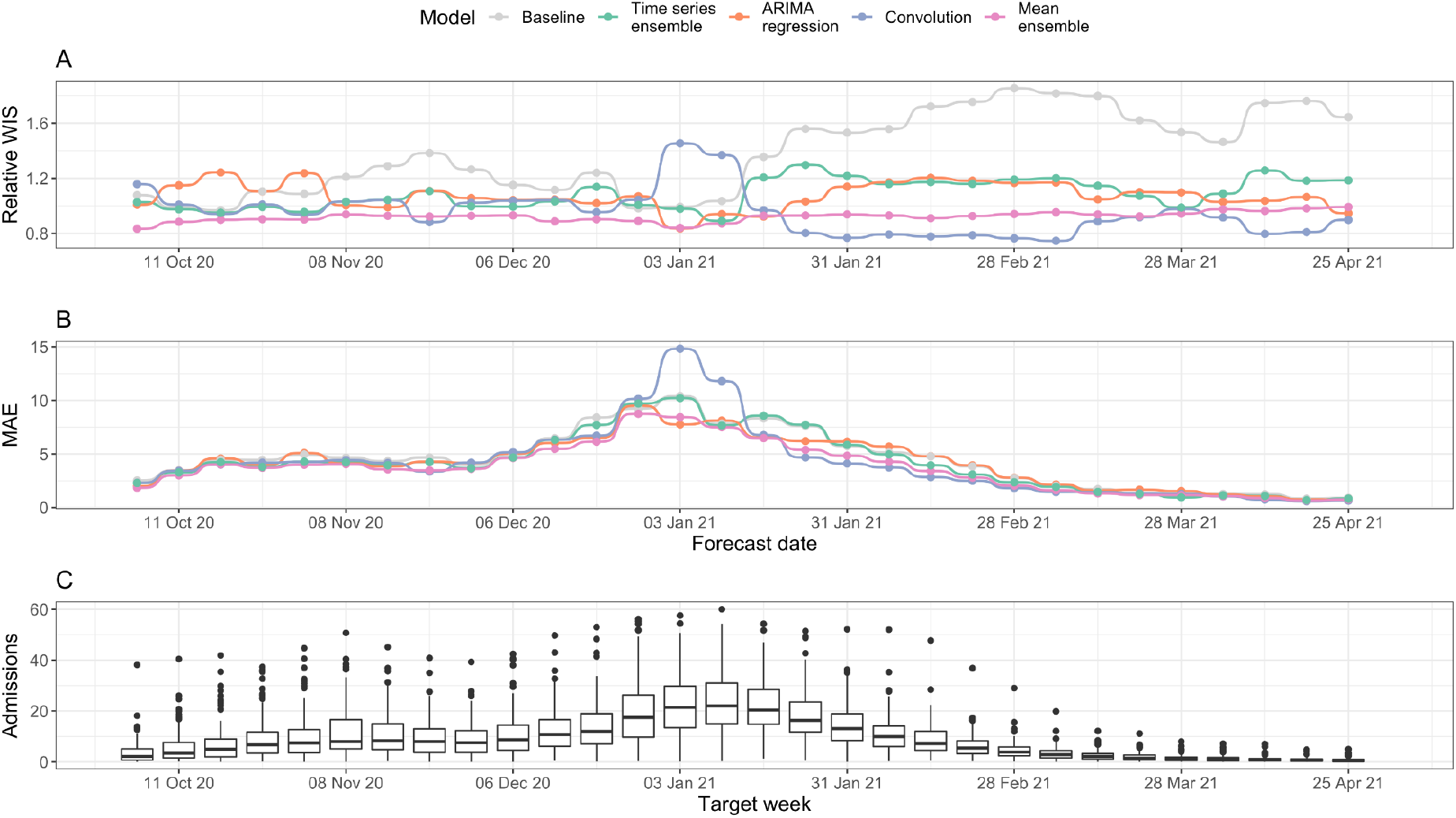
Forecasting accuracy by forecast date (7-day forecast horizon). (A) Relative WIS (rWIS) of the forecasting models for the 30 forecasting dates. Lower rWIS values indicate better forecasts. (B) Mean absolute error of the forecasting models. The mean AE is calculated as the mean AE over all Trusts. (C) Mean daily Trust-level COVID-19 hospital admissions by week, for reference. All panels are for a 7-day forecast horizon; see Additional file 1: Figure S5 for evaluation on a 14-day forecast horizon.

There was less variation in model mean absolute error (MAE) by forecast date (Figure 3B). As expected, the MAE for all models followed the general trend in hospital admissions (Figure 3C), with the exception of forecasts made by the convolution model on 03 and 10 January 2021; for these dates, high rWIS and MAE indicates both poor point and poor quantile forecasts.

For a 14-day horizon, each model performed worse than the baseline on at least one forecast date (Additional file 1: Figure S5A). The decline in performance was especially clear for the convolution model: it only outperformed the baseline on 19/30 forecast dates, and whilst it was the top-ranked model on 12/30 dates, it was also the last-ranked model on 9/30 dates. In particular, the convolution model made noticeably poor forecasts on 08 November, 13 and 27 December 2020, and 03 and 10 January 2021, (Additional file 1: Figure S5A-B).

#### Forecast accuracy by location

For a 7-day horizon, all models outperformed the baseline for the majority of Trusts (Figure 4A): the time series ensemble outperforms the baseline for 125/129 Trusts; the ARIMA regression model for 115/129 Trusts; the convolution model for 118/129 Trusts; and the mean-ensemble for 128/129 Trusts. On average, the mean-ensemble achieved the lowest and most consistent rWIS values (median rWIS = 0.92; IQR = 0.04), compared to median = 1.29, IQR = 0.18 for the baseline. Amongst the individual models, the convolution model had the best median performance (median rWIS = 0.99), but was also the least consistent (IQR = 0.16). The variability in rWIS scores was reflected in the WIS rankings (Figure 4B): the mean-ensemble was best-performing for over half of Trusts (72/129) and first- or second-ranked for almost all Trusts (127/129). The convolution model ranked first through fourth with similar frequency (for 34, 40, 24 and 23/129 Trusts, respectively). Similarly to evaluation by date, we saw less variation in MAE between models by Trust, and a higher MAE in models compared to the baseline more frequently than for the rWIS (Figure 4C).

**Figure 4:**
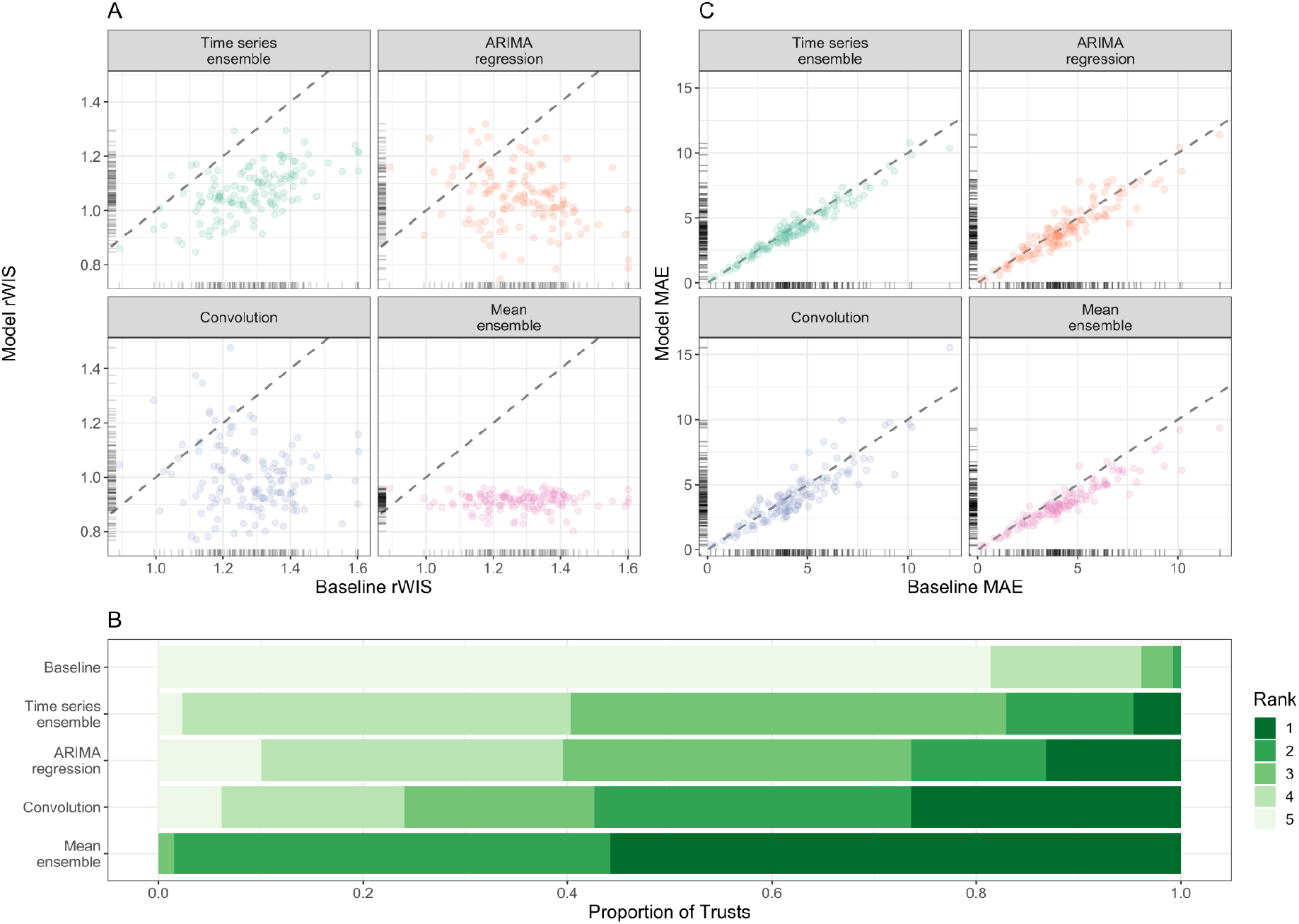
Forecasting accuracy by location (7-day forecast horizon). (A) Relative WIS values of each model (y-axis) compared to the baseline model of no change (x-axis). Ticks on axes show the unilateral distribution of rWIS values. Dashed grey line shows y=x, for reference: a point below the line indicates that the model outperformed the baseline model by rWIS for that Trust. (B) Distribution of WIS rankings across all 129 NHS Trusts; rank 1 is assigned the model with the lowest relative WIS for a given scenario, and rank 5 to the highest relative WIS. (C) Mean absolute error of each model (y-axis) compared to the baseline model (x-axis). Ticks on axes show the unilateral distribution of MAE values. Dashed grey line shows y=x, for reference: a point below the line indicates that the model outperformed the baseline model by MAE for that Trust. All panels are for a 7-day forecast horizon; see Additional file 1: Figure S6 for evaluation on a 14-day forecast horizon.

For a 14-day horizon forecasting accuracy was lower (Additional file 1: Figure S6): the convolution model was particularly badly affected, with a median rWIS equal to that of the baseline model (median rWIS = 1.12) and outperformed the baseline for only 81/129 Trusts. Despite this, the mean-ensemble still performed well: the median rWIS was 0.87 (compared to 1.22 for the baseline), and it was the first- or second-ranked model for 91/129 of Trusts (Additional file 1: Figure S6B).

#### Value of perfect knowledge of future COVID-19 cases

Using future observed COVID-19 cases instead of future forecast cases affects the ARIMA regression model for a forecast horizon of more than 7 days, and the convolution model and mean-ensemble for all forecast horizons. Using future observed cases improved the sWIS by horizon for all affected models (Table S6), especially for the convolution model at a 14-day forecast horizon, where the sWIS decreased by 38% from 1.09 (that is, worse than the baseline model) to 0.67.

When evaluated by forecast date we see a marked improvement in sWIS, but with some variability (Additional file 1: Figure S7A and S7C). The 14-day forecasts made by the ARIMA regression model now outperformed the baseline (sWIS < 1) for 26/30 forecast dates (compared to 22/30 when using forecast future cases; Additional file 1: Figure S7A), with the biggest improvements on forecasts made 13 December 2020 (the start of the spread of the Alpha variant B.1.1.7: sWIS decreased by 26% from 1.33 to 0.99) and 03 January 2021 (just before the third national lockdown: sWIS decreased by 30% from 1.23 to 0.86). The convolution model also saw notable improvements, especially for a 14-day forecast horizon (Additional file 1: Figure S7C). However, for both models there were still forecast dates where they were outperformed by the baseline, indicating that this poor performance was not linked to the case forecasts, but to another aspect of the models.

We also saw an improvement in performance (on average) when we evaluated forecasts by Trust (Additional file 1: Figure S7B and S7D). The 14-day-ahead forecasts made by the ARIMA regression model now outperformed the baseline model for 113/129 Trusts (compared to 102/129 previously; Additional file 1: Figure S7B). The convolution model now outperformed the baseline model for 123/129 Trusts for a 7-day horizon (compared to 116/129), and for 125/129 Trusts for a 14-day horizon (compared to only 81/129 previously; Additional file 1: Figure S7D).

Models using future observed cases as a predictor of future admissions (ARIMA regression, convolution and mean-ensemble) clearly outperformed simple trend-based models, especially at longer forecast horizons (Additional file 1: Figures S8-9). The mean-ensemble performed consistently well across horizons, forecast dates and Trusts, and rarely (if ever) performed worse than the baseline. Conversely, both the ARIMA regression and case-convolution still encountered scenarios where they performed worse than the baseline, warranting further investigation.

## Discussion

This paper systematically evaluates the probabilistic accuracy of individual and ensemble real-time forecasts of Trust-level COVID-19 hospital admissions in England between September 2020 and April 2021. While other COVID-19 forecasting studies evaluate forecasts at the national or regional level [8,21,22], or for small number of local areas (e.g. the city of Austin, Texas, USA [9]; the five health regions of New Mexico, USA [10]; or University College Hospital, London, UK [11], this work evaluates forecast performance over a large number of locations and forecast dates, and explores the usage of aggregate case counts as a predictor of hospital admissions.

We found that all models outperformed the baseline model in almost all scenarios, that is, assuming no change in current admissions was rarely better than including at least a trend. Moreover, models that included cases as a predictor of future admissions generally made better forecasts than purely autoregressive models. However, the utility of cases as a predictor for admissions is limited by the quality of case forecasts: while perfect case forecasts can improve forecasts of admissions, real-time case forecasts are not perfect and can lead to worse forecasts of admissions than simple trend-based models. Unfortunately, making accurate forecasts of COVID-19 cases in a rapidly-evolving epidemic is challenging [23,47], especially in the face of changing local restrictions. The Rt-based case forecasting model used here assumes no change in future Rt, so cannot anticipate sudden changes in transmission, for example due to a change in policy such as lockdowns. Addressing this, and other limitations of the case forecasting model [40], may help to improve admissions forecasts, especially at key moments such as lockdown.

We found that the mean-ensemble model made the most accurate (as measured by median rWIS) and most consistently accurate (as measured by rWIS IQR) forecasts across forecast horizons, forecast dates and Trusts, overcoming the variable performance of the individual models. This is consistent with other COVID-19 forecast evaluation studies [8,15,21,23] and other diseases [25,27,48].

Besides informing situational awareness at a local level, more robust forecasts of hospital admissions can improve forecasts of bed or ICU needs [10,11,14,15], although occupancy forecasts will also depend on patient demographics, patient pathways, ICU requirements and bed availability and length-of-stay distributions [11].

Our framework for forecasting local-level hospital admissions can be applied in other epidemic settings with minimal overheads or used as a baseline to assess other approaches. The models we used are disease-agnostic and only use counts of reported cases and hospital admissions to forecast future admissions. The only context-specific data is the Trust to local authority mapping, used to estimate community pressure of COVID-19 cases on Trusts. In other contexts, this could be replaced with an analogous mapping (either based on admissions data for that disease and/or informed by knowledge of local healthcare-seeking behaviour in that setting), or a mapping based on mobility models of patient flows (e.g. [49,50]). We also note that in other contexts, it may be appropriate to include seasonality in each of the forecasting models.

We found that the prediction interval coverage of the ARIMA regression model was especially low, which inspires a number of areas for future work. One likely reason for this result is that this model uses only the median case forecast, ignoring uncertainty; future work could account for uncertainty of case forecasts (e.g. by using case forecast sample paths as the predictor) and evaluate how this changes the model’s performance. Other reasons for low coverage could be changes over time in the association between cases and admissions, that is, in the CHR or the delay to admission, both of which could occur when the case demographics change [1,12]. Improvements here could allow the lag between cases and admissions to change over time, or to use multiple case predictors at different lags e.g. distributed lag models [51]. However, we also note that these changes carry no guarantee of better forecasting performance: we showed that the case-convolution model (which effectively includes the above adaptations) does not consistently outperform the ARIMA regression model in its current format, especially at longer time horizons.

The mean-ensemble forecast could be further improved in a number of ways, providing many avenues for future work. First, by improving the forecasting accuracy of the existing models, for example by: improving the underlying case forecasts; including additional or more detailed predictors of hospital admissions (e.g. age-stratified cases or mobility). We showed that perfect case forecasts only reduced the WIS of the mean-ensemble by approximately 15% for a 14-day horizon, suggesting efforts would be better spent on identifying better predictors or additional models to include in the ensemble (e.g. other statistical and machine learning models [13,15], or mechanistic models [8]). Other ensemble methods could be considered, such as including a threshold for including models in the ensemble model pool, or making a weighted ensemble based on past performance [8]; however, more complex methods do not guarantee any substantial improvement over a simple mean-ensemble [8,52], and typically require a history of forecast scores to implement. Finally, forecasts may be improved by using a time-varying Trust-UTLA mapping, or by using a mapping with a smaller geographical region (e.g. lower-tier local authorities).

Potential improvements trade off accuracy with data availability (such as: availability in real-time; at a relevant spatial scale and/or across all target locations; whether the data is publicly available) and/or computational power (for additional or more complex forecasting models, or to make reasonable forecasts of additional predictors). During an outbreak, time required to develop and improve forecasting models is limited and in competition with other objectives. When forecasting local-level hospital admissions in epidemic settings, assuming no change in admissions is rarely better than including at least a trend component; including a lagged predictor, such as cases, can further improve forecasting accuracy, but is dependent on making good case forecasts, especially for longer forecast horizons. Using a mean-ensemble overcomes some of the variable performance of individual models and allows us to make more accurate and more consistently accurate forecasts across time and locations.

The models presented here have been used to produce an automated weekly report of hospital forecasts at the NHS Trust level [53] for consideration by policy makers in the UK. Given the minimal data and computational requirements of the models evaluated here, this approach could be used to make early forecasts of local-level healthcare demand, and thus aid situational awareness and capacity planning, in future epidemic or pandemic settings.

## Conclusions

Assuming no change in current admissions is rarely better than including at least a trend. Using confirmed COVID-19 cases as a predictor can improve admissions forecasts in some scenarios, but this is variable and depends on the ability to make consistently good case forecasts. However, ensemble forecasts can make consistently more accurate forecasts across time and locations. Given minimal requirements on data and computation, our admissions forecasting ensemble could be used to anticipate healthcare needs in future epidemic or pandemic settings.

## Data Availability

Fully reproducible data and code are available at https://github.com/epiforecasts/covid19-hospital-activity.

## Abbreviations

AE: absolute error
ARIMA: autoregressive integrated moving average
CHR: Case-hospitalisation rate
COVID-19: coronavirus disease 2019
ETS: exponential smoothing
GLM: generalised linear models
IQR: interquartile range
LAMP: loop-mediated isothermal amplification
MAE: mean absolute error
NHS: National Health Service
PCR: polymerase chain reaction
rWIS: relative weighted interval score
sWIS: scaled weighted interval score
UTLA: upper-tier local authority
WIS: weighted interval score

## Declarations

### Ethics approval and consent to participate

Not applicable.

### Consent for publication

Not applicable.

### Availability of data and materials

All data used in this study are publicly-available. Daily Trust-level COVID-19 hospital admissions are published weekly by NHS England and were accessed via the *covid19*.*nhs*.*data* R package [29]. Daily case reports aggregated by UTLA are published daily on the UK Government dashboard and were accessed via the *covidregionaldata* R package [30]. The Trust-UTLA mapping is available in the R package *covid19*.*nhs*.*data* [29].

### Authors contributions

SM analysed the admissions data and made the forecasts. SM, SA, NB and KS developed the forecasting models. SM drafted the initial manuscript. SM, SA, NB, JM, HG, JH, KS and SF all contributed in interpretation of data, the creation of R packages used in the work, and read, revised and approved the final manuscript. All CMMID COVID-19 Working Group authors contributed in processing, cleaning and interpretation of data, interpreted findings, contributed to the manuscript, and approved the work for publication.

## Acknowledgements

The following authors were part of the CMMID COVID-19 Working Group: Lloyd A C Chapman, Kiesha Prem, Petra Klepac, Thibaut Jombart, Gwenan M Knight, Yalda Jafari, Stefan Flasche, William Waites, Mark Jit, Rosalind M Eggo, C Julian Villabona-Arenas, Timothy W Russell, Graham Medley, W John Edmunds, Nicholas G. Davies, Yang Liu, Stéphane Hué, Oliver Brady, Rachael Pung, Kaja Abbas, Amy Gimma, Paul Mee, Akira Endo, Samuel Clifford, Fiona Yueqian Sun, Ciara V McCarthy, Billy J Quilty, Alicia Rosello, Frank G Sandmann, Rosanna C Barnard, Adam J Kucharski, Simon R Procter, Christopher I Jarvis, Hamish P Gibbs, David Hodgson, Rachel Lowe, Katherine E. Atkins, Mihaly Koltai, Carl A B Pearson, Emilie Finch, Kerry LM Wong, Matthew Quaife, Kathleen O’Reilly, Damien C Tully.

## Additional files

### Additional file 1: supplementary information

**Tab S1**. Summary of fitted ARIMA models. **Tab S2**. Summary of fitted ETS models. **Tab S3**. Forecasting performance (scaled WIS) of time series models. **Fig S1**. Initial estimate of optimal choice of lag between confirmed COVID-19 cases and COVID-19 hospital admissions. **Tab S4**. Scaled WIS of all ARIMA regression models with confirmed COVID-19 cases lagged by d days. **Fig S2**. Daily COVID-19 hospital admissions by England NHS region August 2020 - April 2021. **Fig S3**. Characteristics of acute NHS Trusts in England, August 2020 - April 2021. **Fig S4**. Example of forecasts for Manchester University NHS Foundation Trust. **Tab S5**. Summary of forecasting performance of all forecasting models. **Fig S5**. Forecasting accuracy by forecast date (14-day forecast horizon). **Fig S6**. Forecasting accuracy by location (14-day forecast horizon). **Tab S6**. Value of using observed, vs. forecast, future confirmed COVID-19 cases on forecasting accuracy by forecast horizon. **Fig S7**. Value of using observed, vs. forecast, future confirmed COVID-19 cases on forecasting accuracy by forecast date and location. **Fig S8**. Overall forecasting accuracy of main forecasting models using observed future confirmed COVID-19 cases. **Fig S9**. Forecasting accuracy of main forecasting models using observed future confirmed COVID-19 cases, by forecast date and Trust.

## References

1. Papst I, Li M, Champredon D, Bolker BM, Dushoff J, D Earn DJ. Age-dependence of healthcare interventions for COVID-19 in Ontario, Canada. BMC Public Health. 2021;21: 706.

2. Verity R, Okell LC, Dorigatti I, Winskill P, Whittaker C, Imai N, et al. Estimates of the severity of coronavirus disease 2019: a model-based analysis. Lancet Infect Dis. 2020;20: 669–677.

3. Wilde H, Mellan T, Hawryluk I, Dennis JM, Denaxas S, Pagel C, et al. The association between mechanical ventilator compatible bed occupancy and mortality risk in intensive care patients with COVID-19: a national retrospective cohort study. BMC Med. 2021;19: 213.

4. Carr A, Smith JA, Camaradou J, Prieto-Alhambra D. Growing backlog of planned surgery due to covid-19. BMJ. 2021;372: n339.

5. Camacho A, Kucharski A, Aki-Sawyerr Y, White MA, Flasche S, Baguelin M, et al. Temporal Changes in Ebola Transmission in Sierra Leone and Implications for Control Requirements: a Real-time Modelling Study. PLoS Curr. 2015;7. doi:10.1371/currents.outbreaks.406ae55e83ec0b5193e30856b9235ed2

6. Andronico A, Dorléans F, Fergé J-L, Salje H, Ghawché F, Signate A, et al. Real-Time Assessment of Health-Care Requirements During the Zika Virus Epidemic in Martinique. Am J Epidemiol. 2017;186: 1194–1203.

7. Finger F, Funk S, White K, Siddiqui MR, Edmunds WJ, Kucharski AJ. Real-time analysis of the diphtheria outbreak in forcibly displaced Myanmar nationals in Bangladesh. BMC Med. 2019;17: 58.

8. Funk S, Abbott S, Atkins BD, Baguelin M, Baillie JK, Birrell P, et al. Short-term forecasts to inform the response to the Covid-19 epidemic in the UK. medRxiv; 2020. doi:10.1101/2020.11.11.20220962

9. Arslan N, Sürer O, Morton DP, Yang H, Lachmann M, Woody S, et al. COVID-19 alert stages, healthcare projections and mortality patterns in Austin, Texas, May 2021. 2021. Available: https://covid-19.tacc.utexas.edu/media/filer_public/15/4d/154defa8-9217-478e-a459-8fc4144c61b5/austin_covid_alert_stage_and_mortality_trends_-_ut_-_may_2021.pdf

10. Castro LA, Shelley CD, Osthus D, Michaud I, Mitchell J, Manore CA, et al. How New Mexico Leveraged a COVID-19 Case Forecasting Model to Preemptively Address the Health Care Needs of the State: Quantitative Analysis. JMIR Public Health Surveill. 2021;7: e27888.

11. Leclerc QJ, Fuller NM, Keogh RH, Diaz-Ordaz K, Sekula R, Semple MG, et al. Importance of patient bed pathways and length of stay differences in predicting COVID-19 hospital bed occupancy in England. BMC Health Serv Res. 2021;21: 566.

12. Verhagen MD, Brazel DM, Dowd JB, Kashnitsky I, Mills MC. Forecasting spatial, socioeconomic and demographic variation in COVID-19 health care demand in England and Wales. BMC Med. 2020;18: 203.

13. Vollmer MAC, Glampson B, Mellan T, Mishra S, Mercuri L, Costello C, et al. A unified machine learning approach to time series forecasting applied to demand at emergency departments. BMC Emerg Med. 2021;21: 9.

14. Pagel C, Banks V, Pope C, Whitmore P, Brown K, Goldman A, et al. Development, implementation and evaluation of a tool for forecasting short term demand for beds in an intensive care unit. Operations Research for Health Care. 2017;15: 19–31.

15. Paireau J, Andronico A, Hozé N, Layan M, Crepey P. An ensemble model based on early predictors to forecast COVID-19 healthcare demand in France. 2021. https://hal.sorbonne-universite.fr/METIS-EHESP/pasteur-03149082v1. Accessed 18 January 2022.

16. Alaa A, Qian Z, Rashbass J, Benger J, van der Schaar M. Retrospective cohort study of admission timing and mortality following COVID-19 infection in England. BMJ Open. 2020;10: e042712.

17. Faes C, Abrams S, Van Beckhoven D, Meyfroidt G, Vlieghe E, Hens N, et al. Time between Symptom Onset, Hospitalisation and Recovery or Death: Statistical Analysis of Belgian COVID-19 Patients. Int J Environ Res Public Health. 2020;17. doi:10.3390/ijerph17207560

18. Hyndman RJ, Athanasopoulos G. Forecasting: principles and practice. OTexts; 2018.

19. Ray EL, Reich NG. Prediction of infectious disease epidemics via weighted density ensembles. PLoS Comput Biol. 2018;14: e1005910.

20. Abbott S, Hellewell J, Sherratt K, Gostic K, Hickson J, Badr HS, et al. EpiNow2: Estimate Real-Time Case Counts and Time-Varying Epidemiological Parameters. 2020. doi:10.5281/zenodo.3957489

21. Cramer EY, Lopez VK, Niemi J, George GE, Cegan JC, Dettwiller ID, et al. Evaluation of individual and ensemble probabilistic forecasts of COVID-19 mortality in the US. medRxiv. 2021. doi:10.1101/2021.02.03.21250974

22. Ray EL, Wattanachit N, Niemi J, Kanji AH, House K, Cramer EY, et al. Ensemble forecasts of Coronavirus disease 2019 (COVID-19) in the U.s. medRxiv. 2020. doi:10.1101/2020.08.19.20177493

23. Bracher J, Wolffram D, Deuschel J, Görgen K, Ketterer JL, Ullrich A, et al. A pre-registered short-term forecasting study of COVID-19 in Germany and Poland during the second wave. Nat Commun. 2021;12: 5173.

24. Reich NG, Brooks LC, Fox SJ, Kandula S, McGowan CJ, Moore E, et al. A collaborative multiyear, multimodel assessment of seasonal influenza forecasting in the United States. Proc Natl Acad Sci U S A. 2019;116: 3146–3154.

25. Viboud C, Sun K, Gaffey R, Ajelli M, Fumanelli L, Merler S, et al. The RAPIDD ebola forecasting challenge: Synthesis and lessons learnt. Epidemics. 2018;22: 13–21.

26. Yamana TK, Kandula S, Shaman J. Superensemble forecasts of dengue outbreaks. J R Soc Interface. 2016;13. doi:10.1098/rsif.2016.0410

27. Oidtman RJ, Omodei E, Kraemer MUG, Castañeda-Orjuela CA, Cruz-Rivera E, Misnaza-Castrillón S, et al. Trade-offs between individual and ensemble forecasts of an emerging infectious disease. medRxiv. 2021. doi:10.1101/2021.02.25.21252363

28. NHS Authorities and Trusts. https://www.nhs.uk/ServiceDirectories/Pages/NHSTrustListing.aspx. Accessed 9 Jun 2021.

29. Meakin S, Abbott S, Funk S. covid19.nhs.data: NHS trust level Covid-19 data aggregated to a range of spatial scales. 2021. doi:10.5281/zenodo.4447465

30. Palmer J, Sherratt K, Martin-Nielsen R, Bevan J, Gibbs H, Funk S, et al. covidregionaldata: Subnational data for COVID-19 epidemiology. Journal of Open Source Software. 2021. p. 3290. doi:10.21105/joss.03290

31. Sorensen TA. A method of establishing groups of equal amplitude in plant sociology based on similarity of species content and its application to analyses of the vegetation on Danish commons. Biol Skar. 1948;5: 1–34.

32. Abbott S, Hickson J, Ellis P, Badr HS, Allen J, Munday JD, et al. COVID-19: National and Subnational estimates for the United Kingdom. https://epiforecastsio/covid/posts/national/united-kingdom/. Accessed on 16 October 2020.

33. Hyndman RJ, Khandakar Y. Automatic Time Series Forecasting: The forecast Package for R. J Stat Softw. 2008;27: 1–22.

34. Hyndman R, Athanasopoulos G, Bergmeir C, Caceres G, Chhay L, O’Hara-Wild M, et al. forecast: Forecasting functions for time series and linear models. 2021. Available: https://pkg.robjhyndman.com/forecast/

35. Shaub D, Ellis P. forecastHybrid: Convenient Functions for Ensemble Time Series Forecasts. 2020. Available: https://gitlab.com/dashaub/forecastHybrid

36. Abbott S, Bosse N, DeWitt M, Rau A, Chateigner A, Mareschal S, et al. epiforecasts/EpiSoon: Stable forecasting release. 2020. doi:10.5281/zenodo.3833807

37. Zivot E, Wang J. Modeling Financial Time Series with S-PLUS. 2003rd ed. New York, NY: Springer Science & Business Media; 2003.

38. Lauer SA, Grantz KH, Bi Q, Jones FK, Zheng Q, Meredith HR, et al. The Incubation Period of Coronavirus Disease 2019 (COVID-19) From Publicly Reported Confirmed Cases: Estimation and Application. Ann Intern Med. 2020;172: 577–582.

39. Ganyani T, Kremer C, Chen D, Torneri A, Faes C, Wallinga J, et al. Estimating the generation interval for coronavirus disease (COVID-19) based on symptom onset data, March 2020. Euro Surveill. 2020;25. doi:10.2807/1560-7917.ES.2020.25.17.2000257

40. Abbott S, Hellewell J, Thompson RN, Sherratt K, Gibbs HP, Bosse NI, et al. Estimating the time-varying reproduction number of SARS-CoV-2 using national and subnational case counts. Wellcome Open Res. 2020;5: 112.

41. ONS Estimates of the population for the UK, England and Wales, Scotland and Northern Ireland. 2021. https://www.ons.gov.uk/peoplepopulationandcommunity/populationandmigration/populationestimates/datasets/populationestimatesforukenglandandwalesscotlandandnorthernireland. Accessed 18 January 2022.

42. Gneiting T, Raftery AE. Strictly Proper Scoring Rules, Prediction, and Estimation. J Am Stat Assoc. 2007;102: 359–378.

43. Bracher J, Ray EL, Gneiting T, Reich NG. Evaluating epidemic forecasts in an interval format. PLoS Comput Biol. 2021;17: e1008618.

44. Bosse NI, Abbott S, EpiForecasts, Funk S. scoringutils: Utilities for Scoring and Assessing Predictions. 2020. doi:10.5281/zenodo.4618017

45. NHS COVID-19 Hospital Activity. https://www.england.nhs.uk/statistics/statistical-work-areas/covid-19-hospital-activity/. Accessed 29 September 2021.

46. Vaccinations in England. In: Coronavirus (COVID-19) in the UK. https://coronavirus.data.gov.uk/details/vaccinations?areaType=nation&areaName=England. Accessed 29 Sep 2021.

47. Reich NG, Tibshirani RJ, Ray EL, Rosenfeld R. On the predictability of COVID-19. International Institute of Forecasters. https://forecasters.org/blog/2021/09/28/on-the-predictability-of-covid-19/. Accessed 29 September 2021.

48. Reich NG, McGowan CJ, Yamana TK, Tushar A, Ray EL, Osthus D, et al. Accuracy of real-time multi-model ensemble forecasts for seasonal influenza in the U.S. PLoS Comput Biol. 2019;15: e1007486.

49. Fabbri D, Robone S. The geography of hospital admission in a national health service with patient choice. Health Econ. 2010;19: 1029–1047.

50. Balia S, Brau R, Marrocu E. What drives patient mobility across Italian regions? Evidence from hospital discharge data. Dev Health Econ Public Policy. 2014;12: 133–154.

51. Gasparrini A. Distributed lag linear and non-linear models in R: the package dlnm. J Stat Softw. 2011. Available: https://www.ncbi.nlm.nih.gov/pmc/articles/pmc3191524/

52. Brooks LC, Ray EL, Bien J, Bracher J, Rumack A, Tibshirani RJ, et al. Comparing ensemble approaches for short-term probabilistic covid-19 forecasts in the US. International Institute of Forecasters.https://forecasters.org/blog/2020/10/28/comparing-ensemble-approaches-for-short-term-probabilistic-covid-19-forecasts-in-the-u-s/. Accessed 18 January 2022.

53. Meakin S, Abbott S. epiforecasts/covid19-hospital-activity. 2021. Available: https://github.com/epiforecasts/covid19-hospital-activity

