## Additional file 1 (supplementary information) for "Comparative assessment of methods for short-term forecasts of COVID-19 hospital admissions in England at the local level"

---

**1. Trust-UTLA mapping** **2**

**2. Hospital admissions forecasting models** **2**

2.1. Time series ensemble 2

2.2. ARIMA regression 4

2.3. Case-convolution 6

**3. Case forecasting model** **6**

**4. COVID-19 hospital activity** **8**

**5. Forecast examples** **9**

**6. Forecast evaluation** **9**

6.1. Overall performance 9

6.2. By forecast date 10

6.3. By location 11

**7. Value of perfect knowledge of future COVID-19 cases** **11**

**8. Additional references** **14**

### 1. Trust-UTLA mapping

Confirmed COVID-19 cases are published at the local authority level. To use these data in forecasting models requires estimating a mapping between NHS Trusts and local authorities.

The mapping between NHS Trusts and upper-tier local authorities (UTLAs) is derived from a raw mapping provided by NHS England, based on the Secondary Uses Service (SUS) healthcare data for England. This raw mapping counts the number of COVID-19 hospital discharges of between 01 January 2020 and 30 September 2020 from an NHS site (hospital) to lower-tier local authorities (LTLAs). Site-LTLA pairs are aggregated to Trust-UTLA pairs and we exclude any Trust-UTLA pairs where the number of discharges is less than 10, to preserve anonymity. The mapping between a Trust-UTLA pair is reported as two proportions: the estimated proportion of admissions to a given Trust were admitted from a given UTLA; and the estimated proportion of admissions from a given UTLA that were admitted to a given Trust. The mapping is available in the R package *covid19.nhs.data* [29].

### 2. Hospital admissions forecasting models

#### 2.1. Time series ensemble

We use a mean-ensemble of three simple time series ensemble models: the baseline model (described above), autoregressive integrated moving average (ARIMA) model and exponential smoothing (ETS) model. ARIMA models make forecasts using linear combinations of $p$ past values of the variable of interest and $q$ past forecast errors; if the data is not stationary then one or more additional differencing steps are included. Forecasts made with ETS models are weighted averages of past observed points, where weights are exponentially decreasing; where a trend and/or seasonality is present, exponential smoothing is applied recursively to these components. For further details of the mathematical formulation of the ARIMA and ETS models, we refer the reader to [18].

The time series ensemble model is implemented using the R packages *forecast* [33, 34], *forecastHybrid* [35] and *EpiSoon* [36]: each individual model is fit independently and the final ensemble forecasts are created by taking the mean of the point and interval forecasts from the three models.

A summary of the fitted ARIMA and ETS models is given below (Table S1 and S2). We did not include a day-of-the-week (seasonal) component in the models for which we evaluate forecasts; however, we have also summarised the structure of models where a seasonal component could be included, for reference. Where we allowed a seasonal component to be included, only 726/3,780 (19%) of ARIMA models, and 111/3,780 (3%) of ETS models did include this.

| Parameter | Value | Number of Trust-date pairs (% total) | |
|---|---|---|---|
| | | Non-seasonal | Seasonal |
| p | 0 | 2,861 (76%) | 2,887 (76%) |

| | | | |
|---|---|---|---|
| (number of autoregressive terms) | 1 | 425 (11%) | 420 (11%) |
| | 2 | 291 (8%) | 280 (8%) |
| | 3 | 125 (3%) | 128 (3%) |
| | 4 | 68 (2%) | 58 (2%) |
| | 5 | 10 (<1%) | 7 (<1%) |
| d (number of differencing steps) | 0 | 939 (25%) | 941 (25%) |
| | 1 | 2837 (75%) | 2835 (75%) |
| | 2 | 4 (<1%) | 4 (<1%) |
| q (number of moving average terms) | 0 | 1,439 (38%) | 1,457 (39%) |
| | 1 | 2,055 (54%) | 2,090 (55%) |
| | 2 | 255 (7%) | 204 (5%) |
| | 3 | 30 (1%) | 25 (1%) |
| | 4 | 1 (<1%) | 4 (<1%) |
| Non-zero intercept | TRUE | 930 (25%) | 927 (25%) |
| Non-zero trend | TRUE | 704 (19%) | 713 (19%) |
| Seasonal | TRUE | 0 | 726 (19%) |

**Table S1: Summary of fitted ARIMA models.** Non-seasonal and seasonal models were fitted for each Trust and forecast date. We report the number of Trust-date pairs (3780 total) with values for each of p (the number of autoregressive terms), d (the number of differencing steps) and q (the number of moving average terms), as well as whether an intercept and/or trend was included. We also summarise the number of Trust-date pairs for which an ARIMA model with any seasonal components was fitted.

| Parameter | Value | Number of Trust-date pairs (% total) | |
|---|---|---|---|
| | | Non-seasonal | Seasonal |
| E (Error) | None | 0 | 0 |
| | Additive | 2779 (74%) | 2747 (73%) |
| | Multiplicative | 1001 (26%) | 1033 (27%) |
| T (Trend) | None | 2039 (54%) | 2077 (55%) |
| | Additive | 1741 (46%) | 1703 (45%) |
| | Multiplicative | 0 | 0 |
| S (Seasonal) | None | 3780 (100%) | 3669 (97%) |
| | Additive | 0 | 67 (2%) |

| | | | |
|---|---|---|---|
| | Multiplicative | 0 | 44 (1%) |

**Table S2: Summary of fitted ETS models.** Non-seasonal and seasonal models were fitted for each Trust and forecast date. We report the number of Trust-date pairs (3780 total) where each component was additive, multiplicative, or not included (“None”). Seasonal models were only fit here for comparison and are not included in the forecast evaluation.

Forecasting performance of the three time series models (baseline, ARIMA and ETS) and two mean-ensembles (ARIMA + ETS, and ARIMA + ETS + baseline) by forecast horizon (7 and 14 days) is shown in Table S3.

| Model | Scaled WIS (sWIS) | |
|---|---|---|
| | 7-day horizon | 14-day horizon |
| *Baseline* | *1.00* | *1.00* |
| ARIMA | 0.89 | 0.94 |
| ETS | 0.90 | 0.97 |
| Mean-ensemble: ARIMA + ETS | 0.85 | 0.90 |
| Mean-ensemble: ARIMA + ETS + baseline | 0.84 * | 0.85 * |

**Table S3: Forecasting performance (scaled WIS) of time series models.** Forecasting accuracy for a 7- and 14-day accuracy are shown for three individual models (baseline, ARIMA and ETS), plus two mean ensembles.

#### 2.2. ARIMA regression

In ARIMA regression models, the error term is modelled as an non-seasonal ARIMA process, and therefore allows for autocorrelated error terms. For forecasts of hospital admissions we used Trust-level confirmed COVID-19 cases lagged by $d$ days and estimated through the Trust-UTLA mapping described above. Possible values for the lag $d$ are chosen by considering the Kendall rank correlation coefficient between confirmed COVID-19 cases and hospital admissions at the national and regional (NHS) level between August 2020 and May 2021 (Figure S3). We then made forecasts for a subset of values ( $d = 0, 4, 5, \ldots, 9, 10$), and the optimal value ($d = 7$) chosen as the best-performing model according to the overall WIS rankings (Table S4).

We emphasise that the cross-correlation should not be used as the sole measure by which to determine the optimal lag between cases and admissions, for two reasons. First, optimising for the cross-correlation does not guarantee optimal forecasting performance, such as seen in this instance. Second, this approach may lead to spurious correlation in instances where the two time series are integrated but not co-integrated [37].

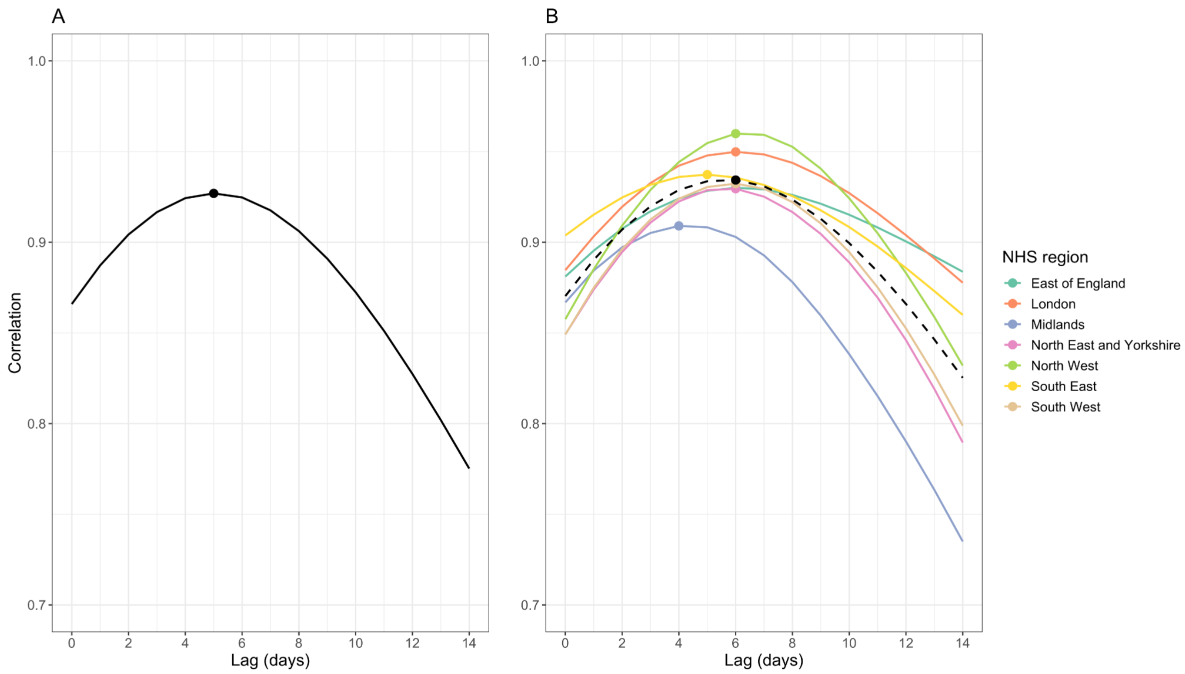


**Figure S1: Initial estimate of optimal choice of lag between confirmed COVID-19 cases and COVID-19 hospital admissions.** Kendall's rank cross-correlation coefficient with a 0 - 14 day lag between confirmed COVID-19 caes and hospital admissions (A) nationally, and (B) by NHS region; dashed line indicates mean correlation over the seven NHS regions. On both panels, the point indicates the lag at which the correlation is maximised: 5 days nationally, and 4-6 days by NHS region.

| Lag *d* (days) | Scaled WIS | |
|---|---|---|
| | 7-day horizon | 14-day horizon |
| 0 | 0.95 | 1.23 |
| 4 | 0.90 | 1.04 |
| 5 | 0.89 | 1.01 |
| 6 | 0.87 | 1.03 |
| 7 | 0.83 * | 0.87 * |
| 8 | 0.86 | 0.88 |
| 9 | 0.89 | 0.88 |
| 10 | 0.89 | 0.95 |

**Table S4: Scaled WIS of all ARIMA regression models with confirmed COVID-19 cases lagged by *d* days.** Scaled WIS values marked with an asterisk (*) indicate the lag which has the lowest (best) score amongst all lags considered.

#### 2.3. Case-convolution

Trust admissions at time $t$ ($A_t$) are assumed to be generated from a negative binomial observation model with mean $\mu_t$ and overdispersion $\phi$. The mean is a convolution of estimated Trust-level COVID-19 cases ($C^*_{t-\tau}$) with the confirmation-to-admission delay distribution ($\xi_\tau$), then scaled by the Trust-level case-hospitalisation ratio ($\alpha$). In mathematical notation:

$$A_t \sim NegBin(\mu_t, \phi)$$

$$\mu_t = \alpha \sum_{\tau=0}^{30} \xi_\tau C^*_{t-\tau} .$$

The parameters of the model were estimated during model fitting with the following weakly informed priors:

$$\alpha \sim N(0.2, 0.1),$$
$$\xi \sim LogNormal(\mu_\xi, \sigma_\xi),$$
$$\mu_\xi \sim N(2.5, 0.5),$$
$$\sigma_\xi \sim N(0.47, 0.25),$$
$$\phi \sim 1/\sqrt{N(0,1)}.$$

We do not include any seasonal effect, as previous work (unpublished) showed that incorrectly including a day-of-the-week effect in the case-convolution model (implemented as a simplex with a Dirichlet prior) led to additional uncertainty and subsequently worse model performance when a day of the week effect could not be identified.

The case-convolution model is fit and forecasts made independently for each date-location pair. Six weeks of data are used for fitting, including a 2-week burn-in period.

### 3. Case forecasting model

Forecasts of COVID-19 cases by UTLA (n = 174) were made via estimates and forecasts of the time-varying effective reproduction number, $R_t$, whilst accounting for uncertainty in the delay distributions. A summary is given below and in [40].

UTLA-level cases at time $t$ were assumed to be generated from a negative binomial observation model with overdispersion $\phi$ and mean $D_t$ (representing the number of cases by date of report at time $t$) scaled by a day-of-the-week effect with an independent parameter for each day of the week ($\omega_{t \bmod 7}$). Cases by date of report are a convolution of the onset-to-report distribution ($\xi_\tau$) and cases by date of onset ($O_{t-\tau}$), which are themselves a convolution of infections ($I_{t-\tau}$) with the incubation period ($\xi_\tau$).

Infections at time $t$ are modelled as a convolution of previous infections with the generation interval distribution ($w_\tau$), scaled by $R_t$. Temporal variation in Rt is controlled by a Gaussian process ($GP$).

In mathematical notation:

$$C_t \sim NegBin(\omega_{t \bmod 7} D_t, \phi)$$

$$D_t = \sum_\tau \xi_\tau O_{t-\tau}$$

$$O_t = \sum_\tau \zeta_\tau I_{t-\tau}$$

$$R_t \sim R_{t-1} \times GP.$$

We assume a log-Normal incubation period with Normal-distributed hyperpriors on the mean ($\mu_\zeta \sim N(5.2, 1.1^2)$) and standard deviation ($\sigma_\zeta \sim N(1.52, 1.1^2)$) [38]. We assume a gamma-distributed generation time $w$ with Normal-distributed hyperpriors on the mean ( $\mu_w \sim N(3.6, 0.7^2)$) and standard deviation ($\sigma_w \sim N(3.1, 0.8^2)$); this is derived from [39], but refit with the above log-Normal distributed incubation period [40]. We also assume a log-Normal onset-to-report delay distribution, again with Normal-distributed hyperpriors on the mean ($\mu_\xi \sim N(0.523, 0.101^2)$) and standard deviation ($\sigma_\xi \sim N(0.945, 0.083^2)$) [40]. We assume $\phi \sim 1/\sqrt{N(0, 1)}$.

Forecasts of UTLA-level COVID-19 cases at time $t + h$ are then made by assuming that $R_{t+h} = R_t$, from which we estimated future infections $I_{t+h}$ and then cases $C_{t+h}$ by the model outlined above. Future changes in contact rates, mobility, or non-pharmaceutical interventions are not accounted for.

### 4. COVID-19 hospital activity

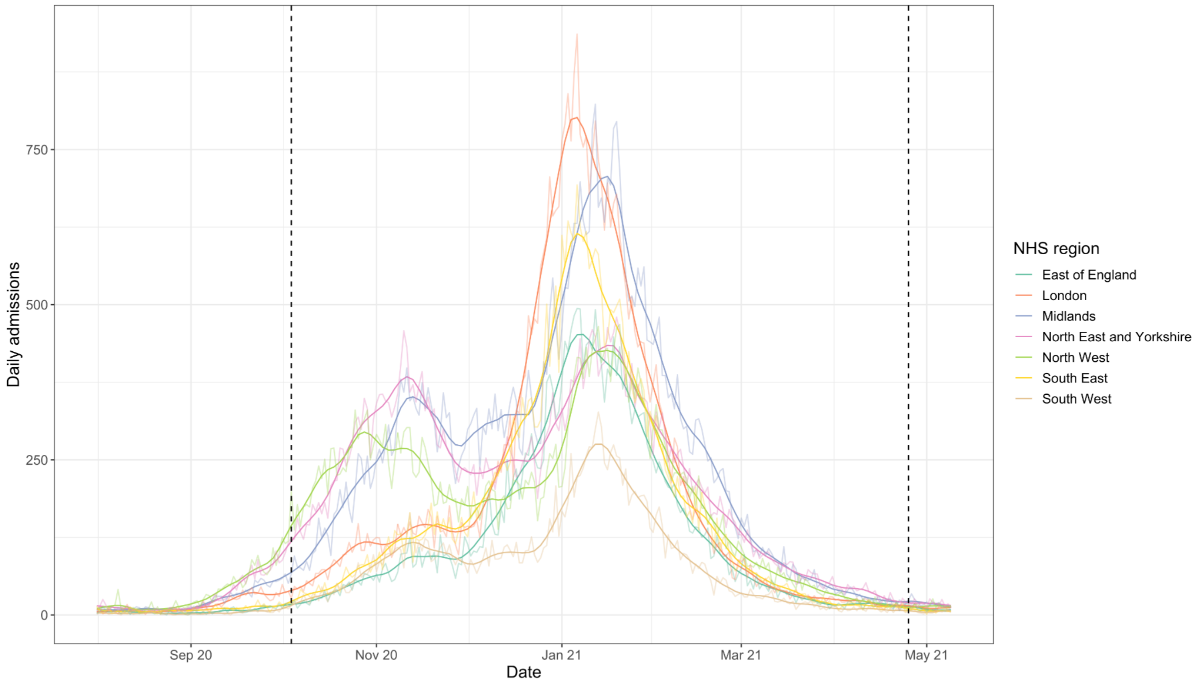


**Figure S2: Daily COVID-19 hospital admissions by England NHS region August 2020 - April 2021.** Raw daily admissions (light) and trend component (dark) of the Seasonal-Trend decomposition using Loess (STL) are both shown for each NHS region (colour); dashed lines denote the first and last forecast date (04 October 2020 and 25 April 2021, respectively).

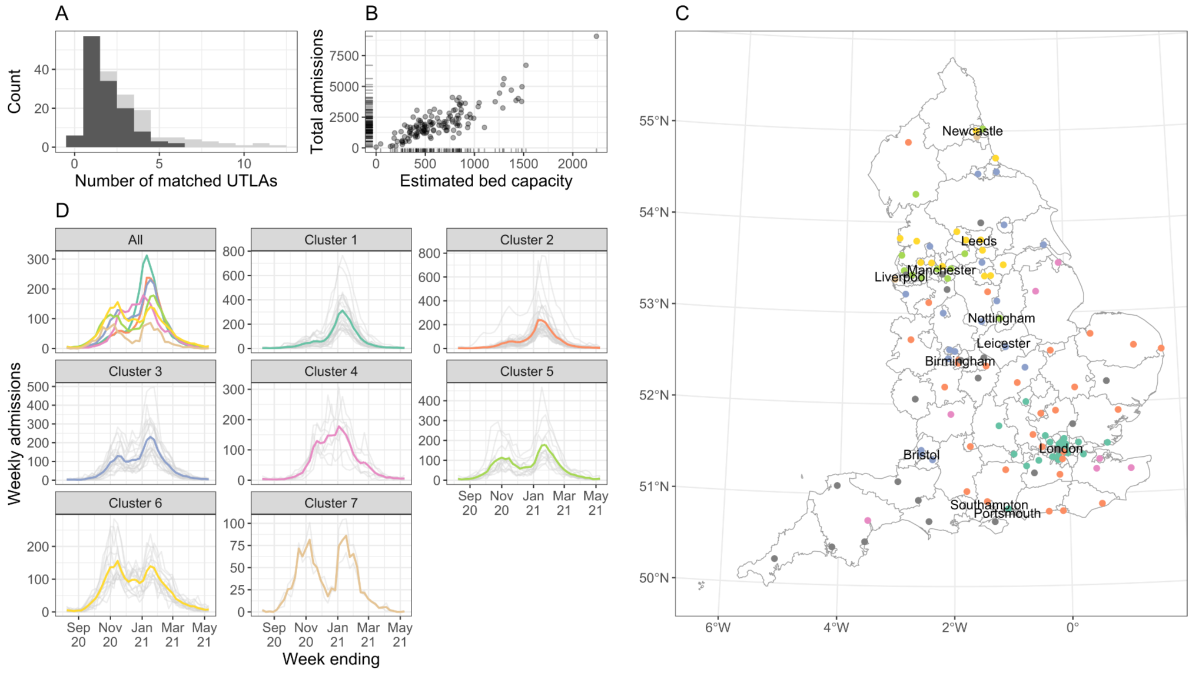


**Figure S3: Characteristics of acute NHS Trusts in England, August 2020 - April 2021.** (A) Distribution of the number of matched UTLAs in the Trust-UTLA mapping, with no threshold (light grey), and with a threshold of 10% on the percentage of admissions from a UTLA (effectively omitting infrequent Trust-UTLA pairs; dark grey). (B) Estimated total bed capacity of Trust (x-axis) vs. total COVID-19 admissions August 2020 - April 2021 (y-axis). Ticks on the axes show the unilateral distribution of estimated bed capacity and total admissions. (C) Locations of acute NHS Trusts in England: colour of point indicates assigned cluster as in panel D, and Trusts with < 1000 total admissions during the study period are shown in grey. Internal boundaries delineate upper-tier local authorities, and main cities are shown for reference. (D) Weekly admissions by cluster: individual Trusts shown in light grey, and the mean of each cluster in colour.

### 5. Forecast examples

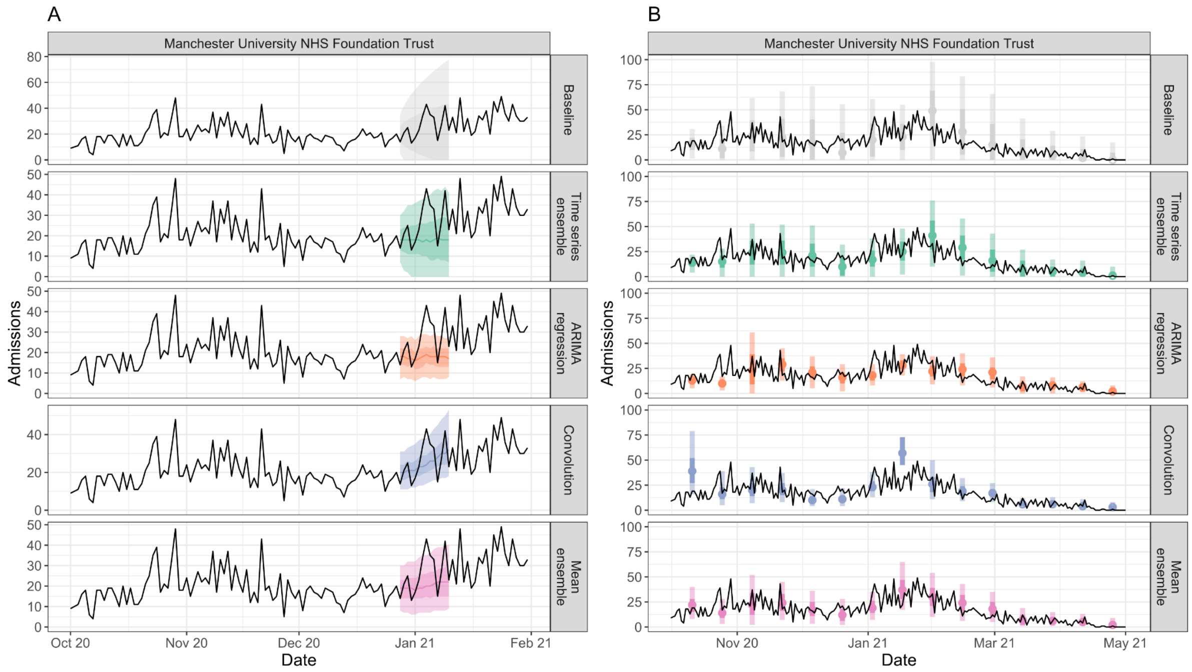


**Figure S4: Example of forecasts for Manchester University NHS Foundation Trust.** (A) Forecasts made on 27 December 2020, showing median forecasts (line), 50% and 90% prediction intervals (dark and light ribbons, respectively) for the full 14-day forecast horizon. (B) Weekly 7-day-ahead forecasts made from 04 October 2020 until 25 April 2021. Points show median forecasts and bars show 50% and 90% prediction intervals (dark and light bars, respectively). In both panels the black line shows observed daily admissions, for reference.

### 6. Forecast evaluation

#### 6.1. Overall performance

| Forecast horizon | Model | 50% emp coverage | 90% emp coverage | Sharp-ness | MAE | rWIS | sWIS |
|---|---|---|---|---|---|---|---|
| 7 days | *Baseline* | *0.70* | *0.82* | *2.15* | *4.46* | *1.26* | *1.00* |
| | Time series ensemble | 0.53 | 0.77 | 1.40 | 4.21 | 1.06 | 0.84 |
| | ARIMA | 0.32 | 0.65 | 0.87 | 4.18 | 1.05 | 0.83 |

| | | | | | | | |
|---|---|---|---|---|---|---|---|
| | regression | | | | | | |
| | Case-convolution | 0.36 | 0.75 | 1.14 | 4.27 | 1.00 | 0.80 |
| | Mean ensemble | 0.45 | 0.78 | 1.13 | 3.68 * | 0.90 | 0.71 * |
| 14 days | *Baseline* | *0.71* | *0.81* | *2.85* | *5.55* | *1.13* | *1.00* |
| | Time series ensemble | 0.54 | 0.75 | 1.76 | 5.51 | 0.97 | 0.85 |
| | ARIMA regression | 0.28 | 0.58 | 0.92 | 5.43 | 0.98 | 0.87 |
| | Case-convolution | 0.33 | 0.72 | 2.07 | 6.95 | 1.23 | 1.09 |
| | Mean ensemble | 0.46 | 0.76 | 1.57 | 4.93 * | 0.86 | 0.76 * |

**Table S5: Summary of forecasting performance of all forecasting models.** Empirical coverage close to the nominal coverage (50% or 90%) is better; lower values of sharpness is better; bias close to zero is better; lower values of mean absolute error (MAE) or relative/scaled WIS (rWIS/sWIS) are better. MAE, rWIS or sWIS marked with an asterisk (*) indicate the model which has the lowest (best) score amongst the models for the given forecast horizon (7 or 14 days) and metric.

#### 6.2. By forecast date

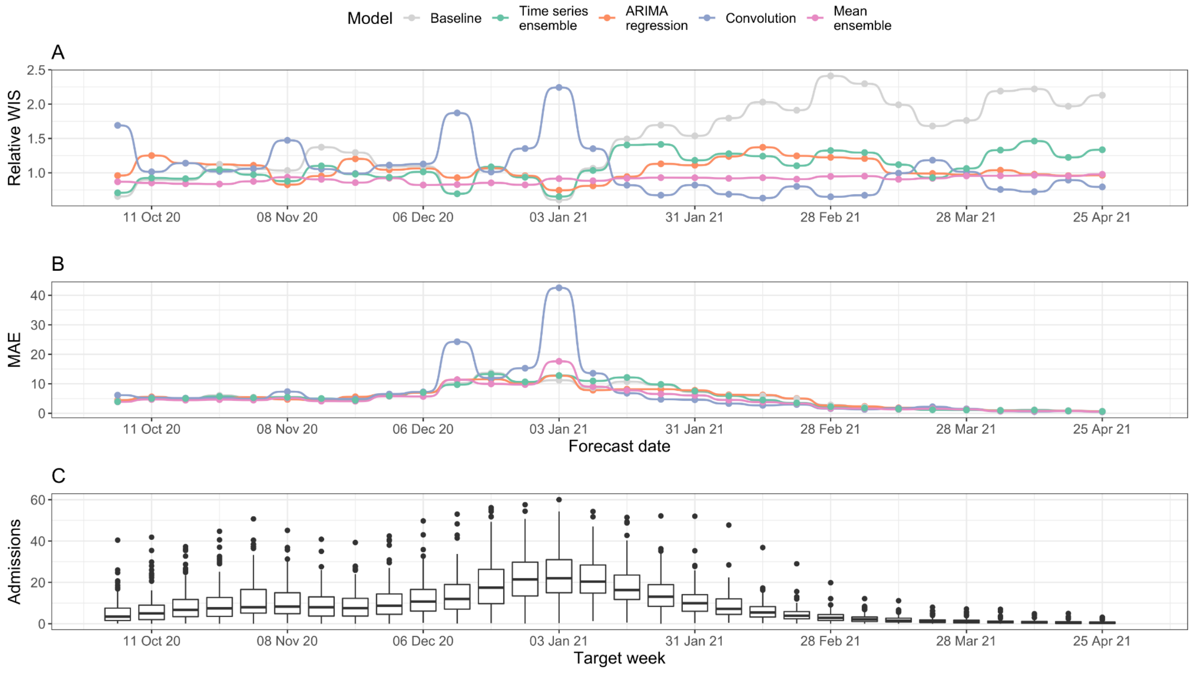


**Figure S5: Forecasting accuracy by forecast date (14-day forecast horizon).** (A) Relative WIS of the main forecasting models by forecast date. (B) Mean absolute error (MAE) of the main forecasting models by forecast date. (C) Mean daily Trust-level COVID-19 hospital admissions by week, for reference. All panels are for a 14-day forecast horizon; see Figure 3 (main text) for evaluation on a 7-day forecast horizon.

#### 6.3. By location

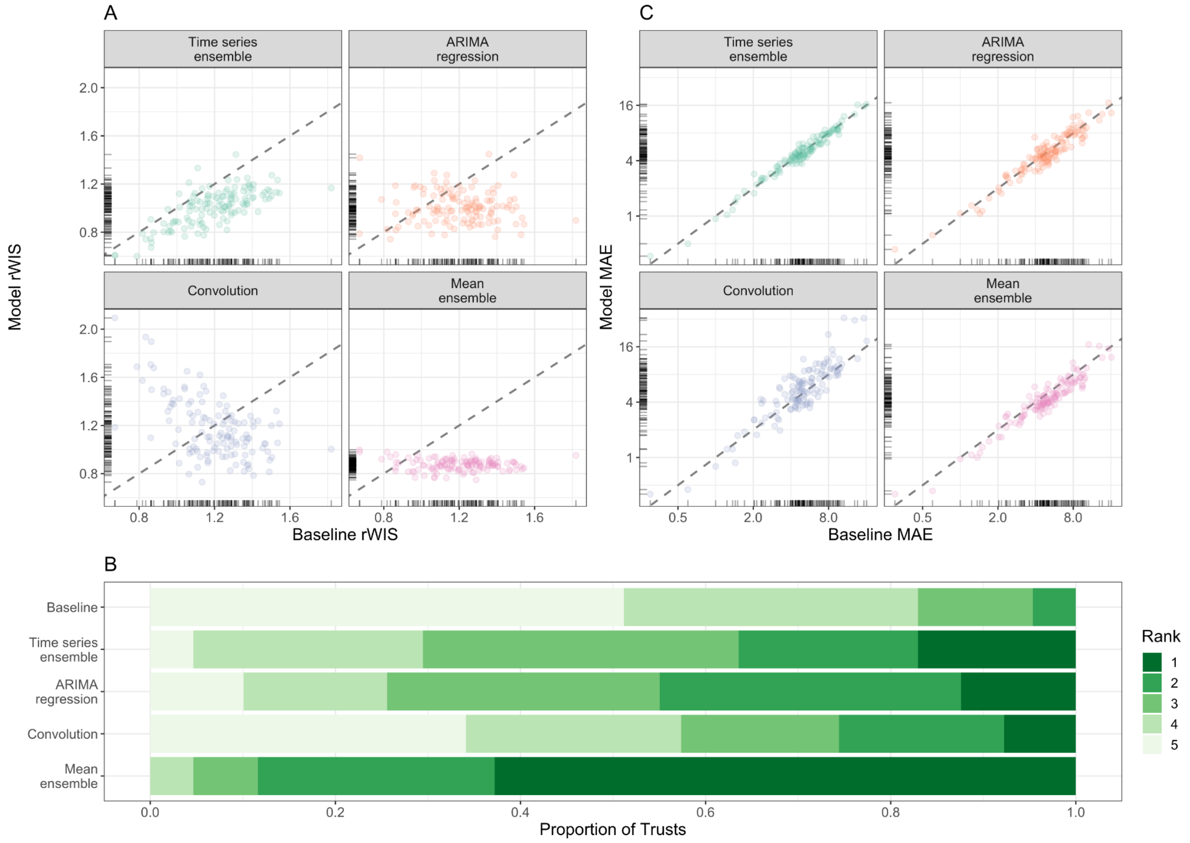


**Figure S6: Forecasting accuracy by location (14-day forecast horizon).** (A) Relative WIS values of each model (y-axis) compared to the baseline model of no change (x-axis). Ticks on axes show the unilateral distribution of rWIS values. Dashed grey line shows y=x, for reference: a point below the line indicates that the model outperformed the baseline model by rWIS for that Trust. (B) Distribution of WIS rankings across all 129 NHS Trusts; rank 1 is assigned the model with the lowest relative WIS for a given scenario, and rank 5 to the highest relative WIS. (C) Mean absolute error of each model (y-axis) compared to the baseline model (x-axis). Ticks on axes show the unilateral distribution of MAE values. Dashed grey line shows y=x, for reference: a point below the line indicates that the model outperformed the baseline model by MAE for that Trust. All panels are for a 14-day forecast horizon; see Figure 4 (main text) for evaluation on a 7-day forecast horizon.

### 7. Value of perfect knowledge of future COVID-19 cases

| Horizon (days) | Model | Scaled WIS | |
|---|---|---|---|
| | | Forecast cases | Observed cases |
| 7 | Time series ensemble | 0.84 | 0.84 |

| | | | |
|---|---|---|---|
| | ARIMA regression | 0.83 | 0.83 |
| | Case-convolution * | 0.80 | 0.76 |
| | Mean ensemble * | 0.71 | 0.71 |
| 14 | Time series ensemble | 0.85 | 0.85 |
| | ARIMA regression * | 0.87 | 0.80 |
| | Case-convolution * | 1.09 | 0.67 |
| | Mean ensemble * | 0.76 | 0.65 |

**Table S6: Value of using observed, vs. forecast, future confirmed COVID-19 cases on forecasting accuracy by forecast horizon.** Models marked with an asterisk (*) denote models that use forecast future COVID-19 cases as a predictor of future admissions at that horizon, and so we would expect to be affected by the change to future observed cases.

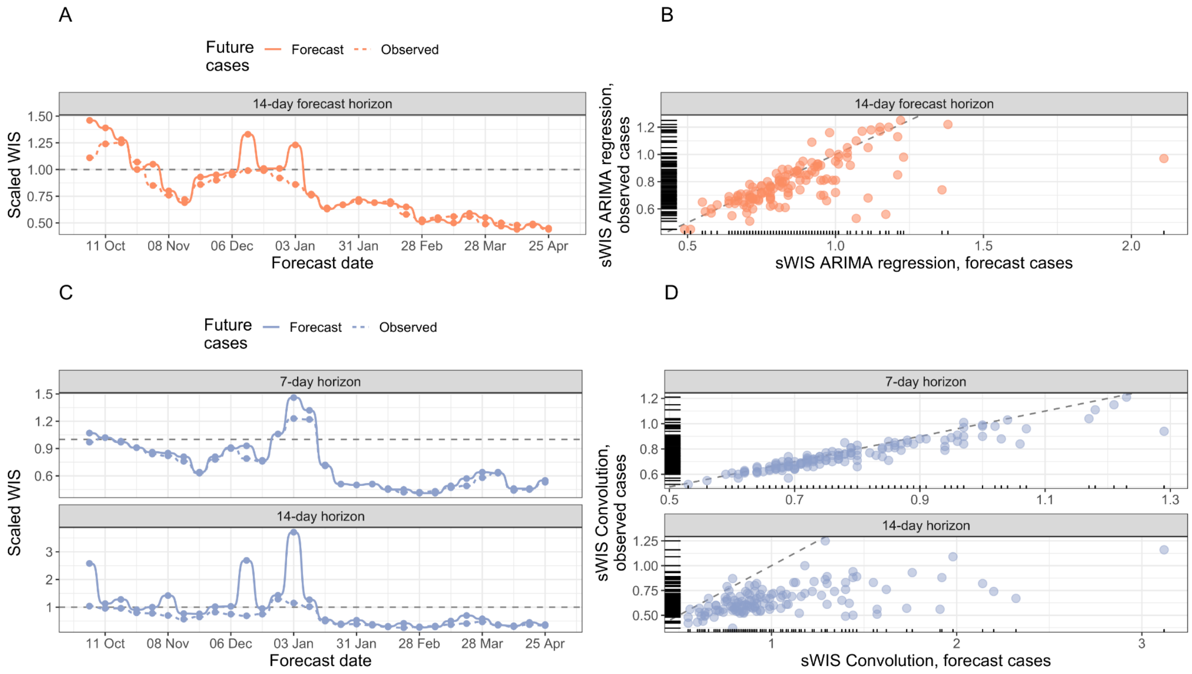


**Figure S7: Value of using observed, vs. forecast, future confirmed COVID-19 cases on forecasting accuracy by forecast date and location.** (A) Scaled WIS (sWIS) values by forecast date for 14-day-ahead forecasts generated by the ARIMA regression model, using observed (solid line) and forecast (dashed line) COVID-19 cases. (B) sWIS values by Trust for 14-day-ahead forecasts generated by the ARIMA regression model. (C) sWIS values by forecast date for 7- and 14-day-ahead forecasts generated by the convolution model, using observed (solid line) and forecast (dashed line) COVID-19 cases; the model using observed cases outperforms the original model for all forecast dates. (D) rWIS values by Trust for 7- and 14-day-ahead forecasts generated by the convolution model. In panels A and C, the dashed line shows y = x, for reference; values below the line indicate an improvement in forecasting performance. In panels B and D, the dashed line shows the scaled WIS of the baseline, for reference. sWIS is shown on the log2 scale in all panels.

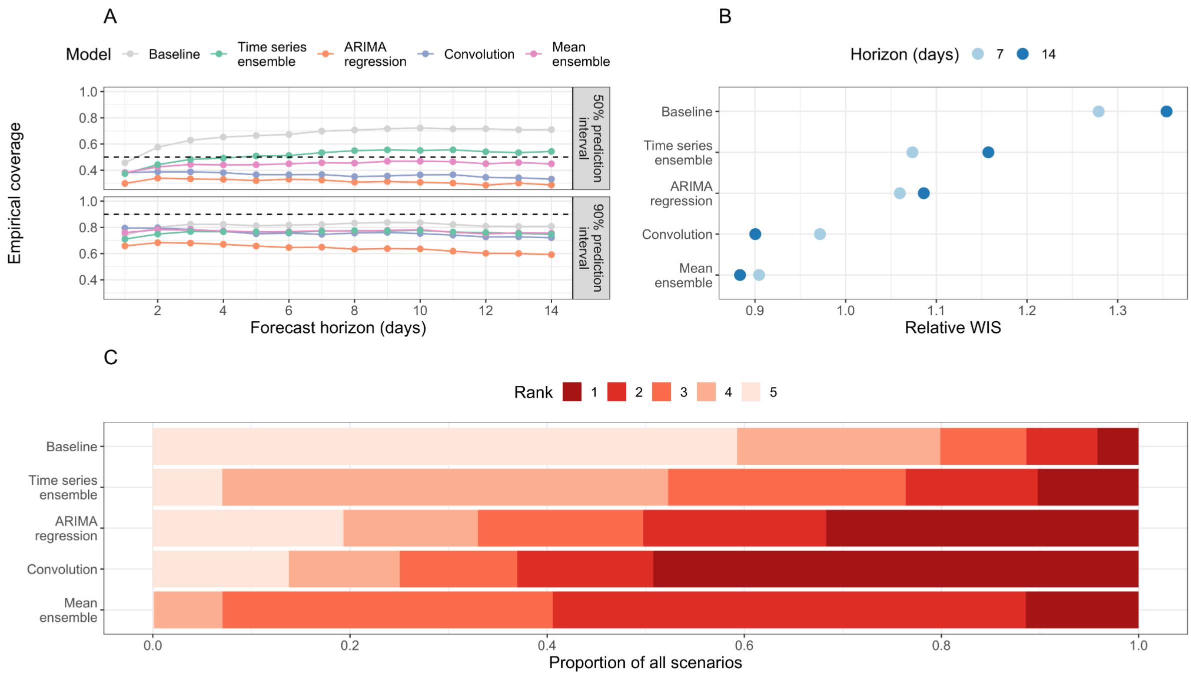


**Figure S8: Overall forecasting accuracy of main forecasting models using observed future confirmed COVID-19 cases.** (A) Empirical coverage of 50% and 90% prediction intervals for 1-14 days forecast horizon. The dashed line indicates the target coverage level (50% or 90%). (B) Relative weighted interval score (rWIS) by forecast horizon (7 and 14 days). (C) Distribution of WIS rankings across all 7,701 targets; for each target, rank 1 is assigned to the model with the lowest relative WIS (rWIS) and rank 5 to the model with the highest rWIS.

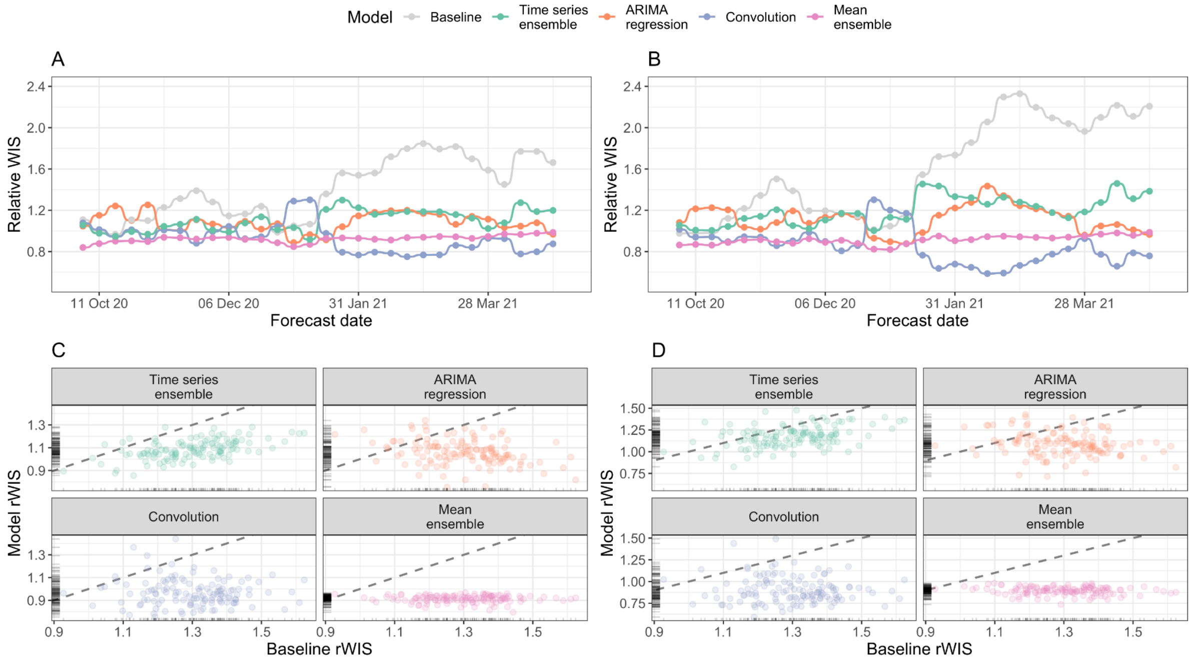


**Figure S9: Forecasting accuracy of main forecasting models using observed future confirmed COVID-19 cases, by forecast date and Trust.** (A - B) Relative WIS (rWIS) of the main forecasting models for a (A) 7-day and (B) 14-day horizon. (C - D) rWIS values (y-axis) compared to the baseline model (x-axis). Ticks on axes show the marginal distribution of rWIS values. Dashed grey line shows y=x, for reference: a point below the line indicates that the model outperformed the baseline model for that Trust. Shown for a (C) 7-day and (D) 14-day horizon.
